# Measuring Mental Health in a Cost-of-Living Crisis: a rapid review

**DOI:** 10.1101/2023.07.24.23293078

**Authors:** Clare England, David Jarrom, Jenni Washington, Elise Hasler, Leona Batten, Ruth Lewis, Rhiannon Tudor Edwards, Jacob Davies, Brendan Collins, Alison Cooper, Adrian Edwards

**Affiliations:** Health Technology Wales, United Kingdom; Health and Care Research Wales Evidence Centre, Bangor University, United Kingdom; Science Evidence and Advice Division, Welsh Government; Health and Care Research Wales Evidence Centre, Cardiff University, United Kingdom

## Abstract

Since 2021 the UK has experienced a sharp rise in inflation. For many, wages and welfare payments have not kept up with rising costs, leading to a cost of living crisis. There is evidence indicating that economic crises are damaging to population mental health and that some groups are particularly vulnerable.

The review aims to 1. Identify and appraise available population-level measures and methods for assessing the impact on mental health of any public health response to the cost-of-living crisis and 2. Review the appropriateness of the measures for specific, vulnerable populations.

**Study designs and mental health measurement tools:** These included secondary analyses of existing data, household panel surveys, repeated cross-sectional surveys; or used routine clinical data including medical records, prescribing data, or were ecological time-series studies using national or regional suicide death rates. 12 validated mental health measurement tools were identified. Four validated mental health measurement tools are embedded into UK population-level surveys.

**Vulnerable groups:** 11 mental health measurement tools were used to identify population sub-groups whose mental health was most likely to be affected by an economic crisis. There is evidence that the mental health measurement tools and methods are suitable for measuring mental health in people with different socio-economic and financial situations. It was not possible to determine whether the methods and tools effectively captured data from people from minority ethnic groups.

**Policy and practice implications:** Many UK population-level surveys, include validated mental health tools and questions about financial security, providing data that can be used to explore population mental health. A quasi-experimental study design, using data from a household panel could be suitable for measuring the mental health impact of a specific public health initiative to tackle cost of living pressures. Reports and studies using population-level surveys or medical records should present data on ethnicity and, where possible, plan to stratify analyses by ethnicity.

**Economic considerations:** Poorer households are more exposed to inflationary pressures. In the lead up to the cost of living crisis, Wales had the highest proportion of working age adults and pensioners in relative income poverty out of the UK nations. 28% of children in Wales were living in relative poverty. Given that over half of all mental health problems start by age 14 (and 75% by age 18) and poverty being a risk factor for psychological illnesses, there is likely to be a long shadow of mental health continuing into future generations stemming from the cost-of-living crisis. Mental Health problems cost the Welsh economy 4.8 billion (UK pounds) per annum. In a recent survey of Welsh participants, 60% of respondents agreed that rising costs of living negatively affected their quality of life.

**Funding statement:** Health Technology Wales was funded for this work by the Health and Care Research Wales Evidence Centre, itself funded by Health and Care Research Wales on behalf of Welsh Government

**EXECUTIVE SUMMARY:** *What is a Rapid Review?:* Our rapid reviews use a variation of the systematic review approach, abbreviating or omitting some components to generate the evidence to inform stakeholders promptly whilst maintaining attention to bias.

*Who is this summary for?:* The intended audience are stakeholders needing to measure mental health outcomes who are seeking evidence for appropriate methods and tools, which are applicable to the UK or relating to Wales.

*Background / Aim of Rapid Review:* Since 2021, the UK has experienced a sharp rise in inflation. For most people, wages and welfare payments have not kept up with rising costs, leading to a cost-of-living crisis. There is evidence drawn from longitudinal epidemiological studies indicating that economic crises are damaging to population mental health and that some groups are particularly vulnerable. Consequently, public health responses to the cost-of-living crisis should be able to assess the impact of the policies on mental health. The aims of the review are to 1) identify and appraise available population-level measures and methods for assessing the impact on mental health of any public health response to the cost-of-living crisis and 2) review the appropriateness of the measures for specific, vulnerable populations.

*Key Findings:* Four systematic reviews, one scoping review, one clinical guidance, one rapid review, nine peer-reviewed primary studies and two reports from grey literature were included. Most evidence come from before and after the 2008/2009 economic crisis, which is also called the Great Recession. Study designs and mental health measurement tools

▪ Study designs included secondary analyses of existing data drawn from national or regional cohort studies, household panel surveys, repeated cross-sectional surveys; or used routine clinical data including medical records, prescribing data, or were ecological time-series studies using national or regional suicide death rates.
▪ Two quasi-experimental studies used data from a household panel survey to examine the impact of the introduction of specific welfare policies on mental health.
▪ Seven UK studies used data from the “Understanding Society: The UK Household Longitudinal Study” (UKHLS), one used the “Scottish Longitudinal Study” (SLS), one used the “Welsh Health Survey” (now “National Survey for Wales”), and one used the Office for National Statistics (ONS) “Opinions and Lifestyle Survey”.
▪ Twelve brief validated mental health measurement tools, which were self-administered, or administered by an interviewer, were identified (section 6, table 4).
▪ Four validated mental health measurement tools are embedded into UK population-level surveys. The four tools are: 12-item General Health Questionnaire (used in UKHLS); Short form 12 Mental Health Component Summary (used in UKHLS); Warwick-Edinburgh Mental Well-being Scale (used in UKHLS and the “National Survey for Wales”) and Patient Health Questionnaire depression scale (used in the “Opinions and Lifestyle Survey”) Vulnerable groups

▪ Eleven of the twelve mental health measurement tools were used to identify population sub-groups whose mental health was most likely to be affected by an economic crisis.
▪ The reviews and studies did not comment on the suitability of the mental health measurement tools for measuring mental health among vulnerable groups.
▪ There is evidence that the identified mental health measurement tools and methods are suitable for measuring mental health in people with different socio-economic and financial situations, including those who are financially insecure and from men and women and from people of different ages.
▪ It was not possible to determine whether the identified methods and tools effectively captured data from people from minority ethnic groups. Policy and practice implications

▪ The UK has many population-level surveys, which include validated mental health tools and questions about financial security, providing rich data that can be used to explore the mental health of the population.
▪ A quasi-experimental study design, using data from a household panel survey such as the UKHLS, could be suitable for measuring the mental health impact of a specific public health initiative to tackle cost-of-living pressures, and which has a clear roll-out date.
▪ Reports and studies using population-level surveys or medical records should present data on ethnicity and, where possible, plan to stratify analyses by ethnicity. Economic considerations

▪ The impacts of the cost of living crisis have not been felt equally. Poorer households are more exposed to inflationary pressures as they spend a greater proportion of their income on items such as food and energy that have seen considerable inflation.
▪ In the lead up to the cost of living crisis, Wales had the highest proportion of working age adults (21%) and pensioners (18%) in relative income poverty out of the UK nations. 28% of children in Wales were living in relative poverty. Given that over half of all mental health problems start by age 14 (and 75% by age 18) and poverty being a known risk factor for psychological illnesses, there is likely to be a long shadow of mental health continuing into future generations stemming from the cost of living crisis.
▪ Mental Health problems cost the Welsh economy £4.8 billion per annum.
▪ In a survey of 2,000 Welsh participants covering the period November 2022 to January 2023, 60% of respondents agreed that rising costs of living negatively affected their quality of life (25% strongly agreed). 87% reported ‘worrying’ around the cost of living, with 38% reporting ‘worrying a lot’.

## 1. BACKGROUND

### 1.1 Who is this review for

This Rapid Review was conducted as part of the ‘Health and Care Research Wales Evidence Centre’ Work Programme. The above question was suggested by the Science, Evidence and Advice (SEA) Division, Welsh Government. The intended audience are stakeholders needing to measure mental health outcomes who are seeking evidence for the appropriate methods and tools which are applicable to the UK or relating to Wales.

### 1.2 Background and purpose of this review

Since 2021 the UK has experienced a sharp rise in inflation. For most people, wages and welfare payments have not kept up with the rate of inflation, leading to a cost-of-living crisis. Energy, food, fuel, and housing have become less affordable, which leads to adverse short and long-term health outcomes (Roberts et al. 2022). There is evidence from past economic crises, such as the 2008/2009 global financial crisis which is also known as The Great Recession, that economic crises and increases in the cost-of-living have a detrimental effect on mental health across the population and that vulnerable populations are disproportionately affected (Guerra & Eboreime 2021). In response, national and regional public health interventions may be put into place to mitigate against the effects of a cost-of-living crisis. It is important for policymakers to be able to measure the impact of any public health measure on mental health.

This rapid review answers the following questions:

- What methods and tools are available and appropriate for monitoring the impact of the cost-of-living crisis on mental health?
- How is it best to do this for specific, vulnerable populations?

The aims of the review are to 1) identify and appraise available population-level measures for assessing the impact on mental health of any public health response to the cost-of-living crisis and 2) review the appropriateness of the measures for specific, vulnerable populations.

The purpose of this review is not to summarise the findings of included articles (i.e., impact of the economic crises on mental health or outcome of any public health interventions), or to identify how often different methods of measurement are used to evaluate the effect of economic crises on mental health, but to provide a summary of the available evaluation methods, and the comments on study design.

## 2. RESULTS

### 2.1 Overview of the Evidence Base

We searched for published articles that evaluated the impact of economic crises on population mental health, that were longitudinal in design and that used validated tools or other clear methods to measure mental health outcomes. Relevant methods were identified from secondary evidence published from 1970 to 2023 and from primary studies published from 2021 to 2023 that were not included in the reviews. We identified four systematic reviews (Frasquilho et al. 2016, Glonti et al. 2015, Saez et al. 2019, Silva et al. 2018), one scoping review (Guerra & Eboreime 2021) one clinical guideline (Martin-Carrasco et al. 2016) and one rapid review from grey literature (Preece & Bimpson 2019) where the majority of included studies met our inclusion criteria. The reviews included a total of 322 different identifiable studies, of which 52 were included in more than one review (although Martin-Carrasco reported using evidence from a total of 354 studies, but detailed data was only extracted from 69). In addition, we identified nine further peer-reviewed primary studies (Alvarez-Galvez et al. 2021, Aretz 2022, Cherrie et al. 2021, Clair & Baker 2022, Curtis et al. 2021, Kim et al. 2022, Saville 2021, Thomson et al. 2022, Wickham et al. 2020) and two reports from grey literature (Clark & Wenham 2022, Office for National Statistics 2022).

Full details of inclusion and exclusion criteria and the screening of articles can be found in section 5 A detailed summary of the methods used in each study, and an overview of identified mental health measurement tools are available in section 6 (tables 3a-3c and table 4).

### 2.2 Summary of the Evidence

#### 2.2.1 Characteristics of included studies

One systematic review (Saez et al. 2019) was specific to Spain, and one rapid review (Preece & Bimpson 2019) had a Welsh focus, although evidence was drawn from other countries. All other reviews did not have limitations on countries but over half of included studies were conducted in Europe. Four reviews (Frasquilho et al. 2016, Glonti et al. 2015, Guerra & Eboreime 2021, Silva et al. 2018) included studies examining the effects of any economic or financial crisis on mental health; three (Martin-Carrasco et al. 2016, Saez et al. 2019, Preece & Bimpson 2019) were limited to the effects of Great Recession.

Seven of the additional primary studies (Cherrie et al. 2021, Clair & Baker 2022, Curtis et al. 2021, Kim et al. 2022, Saville 2021, Thomson et al. 2022, Wickham et al. 2020) were conducted in the UK. The remaining two primary studies were in other European countries (Alvarez-Galvez et al. 2021, Aretz 2022). All nine primary studies were limited to the effects of the Great Recession. The two grey literature reports (Clark & Wenham 2022, Office for National Statistics 2022) were UK focussed and specific to the 2021/2022 cost of living crisis.

#### 2.2.2 Study design and sources of data (table 1)

**Table 1.**
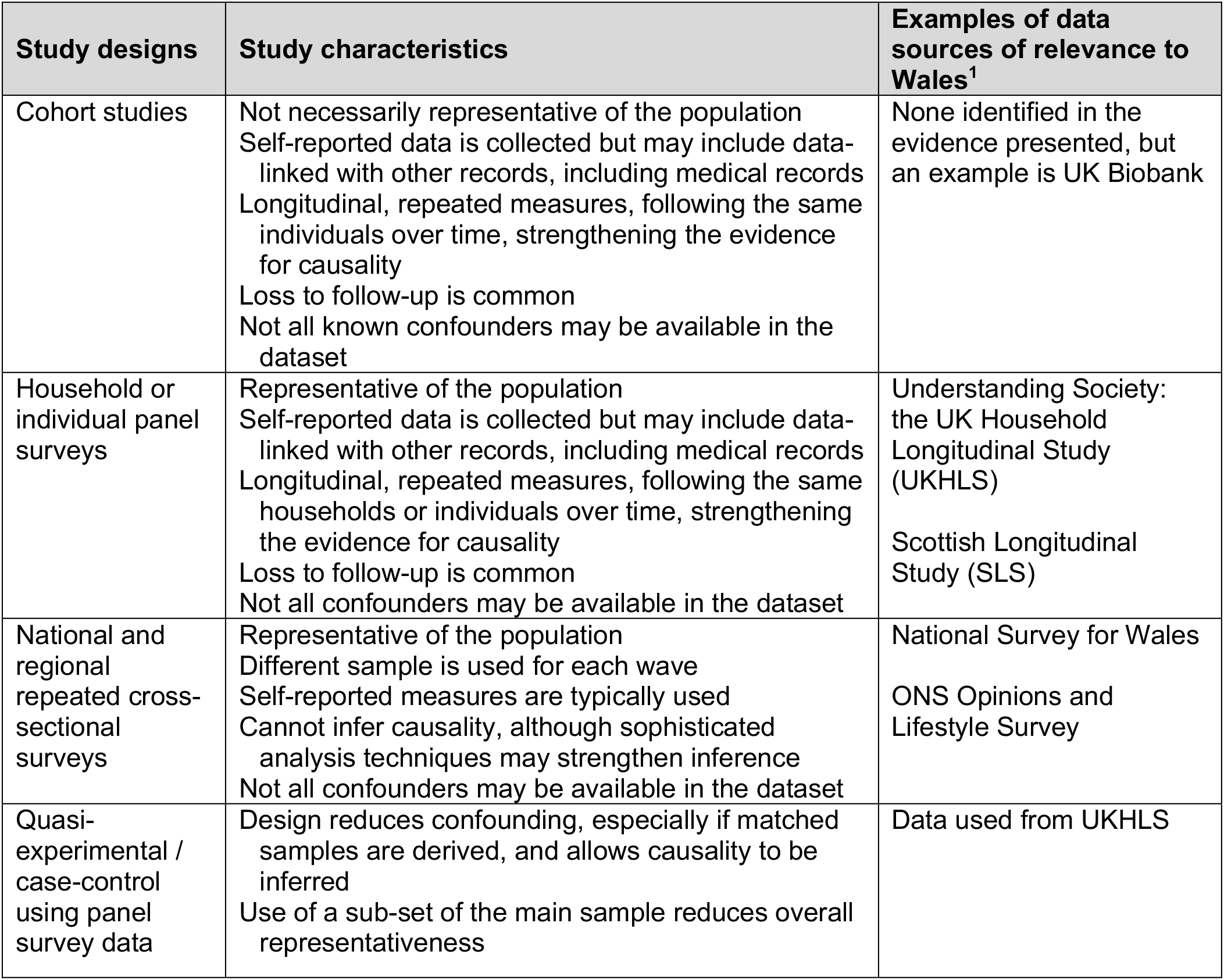
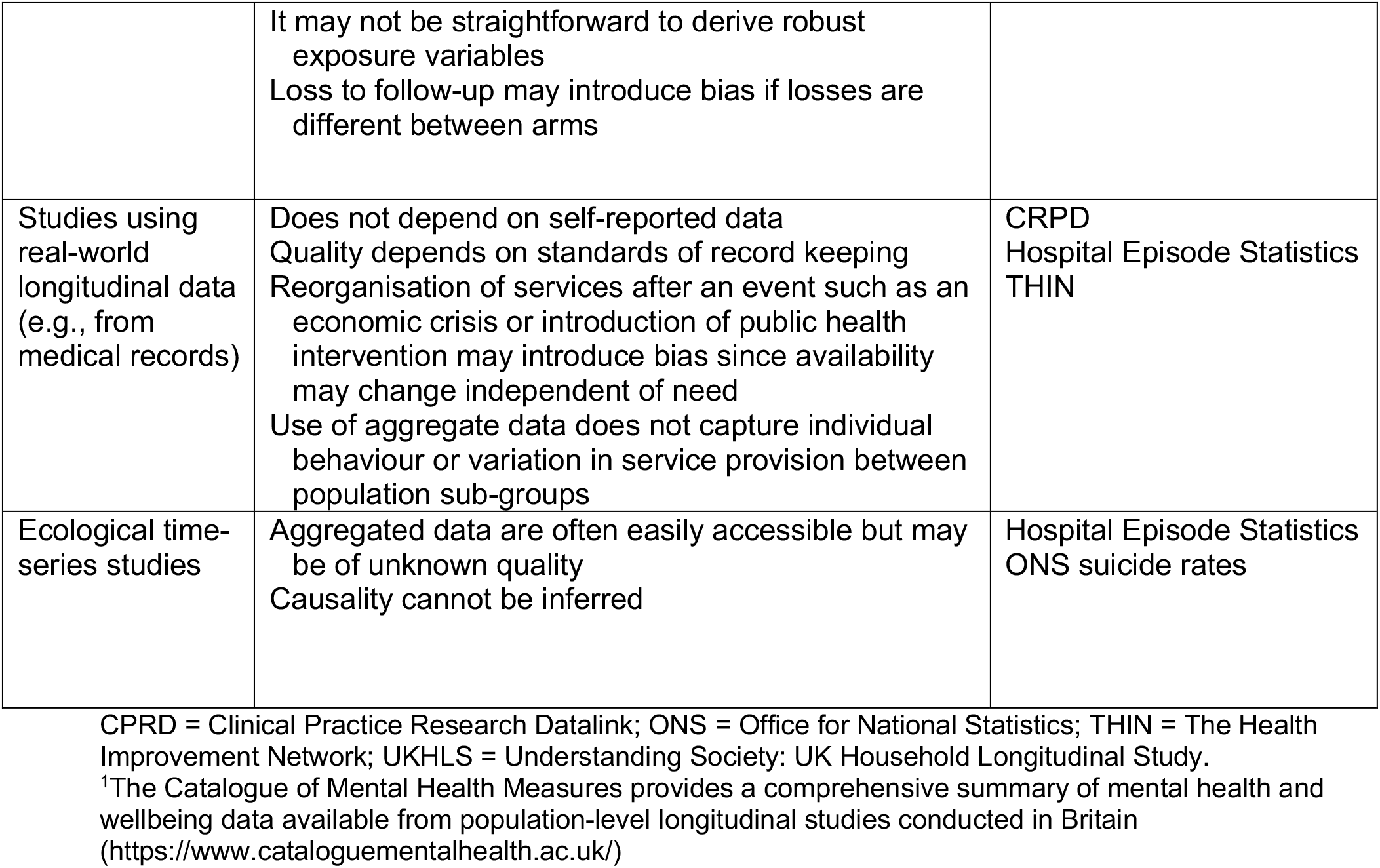
Study designs and main characteristics.

| Study designs | Study characteristics | Examples of data sources of relevance to Wales<sup>1</sup> |
| --- | --- | --- |
| Cohort studies | Not necessarily representative of the population<br>Self-reported data is collected but may include data-linked with other records, including medical records<br>Longitudinal, repeated measures, following the same individuals over time, strengthening the evidence for causality<br>Loss to follow-up is common<br>Not all known confounders may be available in the dataset | None identified in the evidence presented, but an example is UK Biobank |
| Household or individual panel surveys | Representative of the population<br>Self-reported data is collected but may include data-linked with other records, including medical records<br>Longitudinal, repeated measures, following the same households or individuals over time, strengthening the evidence for causality<br>Loss to follow-up is common<br>Not all confounders may be available in the dataset | Understanding Society: the UK Household Longitudinal Study (UKHLS)<br>Scottish Longitudinal Study (SLS) |
| National and regional repeated cross-sectional surveys | Representative of the population<br>Different sample is used for each wave<br>Self-reported measures are typically used<br>Cannot infer causality, although sophisticated analysis techniques may strengthen inference<br>Not all confounders may be available in the dataset | National Survey for Wales<br>ONS Opinions and Lifestyle Survey |
| Quasi-experimental / case-control using panel survey data | Design reduces confounding, especially if matched samples are derived, and allows causality to be inferred<br>Use of a sub-set of the main sample reduces overall representativeness | Data used from UKHLS |
|  | It may not be straightforward to derive robust exposure variables<br>Loss to follow-up may introduce bias if losses are different between arms |  |
| Studies using real-world longitudinal data (e.g., from medical records) | Does not depend on self-reported data<br>Quality depends on standards of record keeping<br>Reorganisation of services after an event such as an economic crisis or introduction of public health intervention may introduce bias since availability may change independent of need<br>Use of aggregate data does not capture individual behaviour or variation in service provision between population sub-groups | CRPD<br>Hospital Episode Statistics<br>THIN |
| Ecological time-series studies | Aggregated data are often easily accessible but may be of unknown quality<br>Causality cannot be inferred | Hospital Episode Statistics<br>ONS suicide rates |

CPRD = Clinical Practice Research Datalink; ONS = Office for National Statistics; THIN = The Health Improvement Network; UKHLS = Understanding Society: UK Household Longitudinal Study.
<sup>1</sup>The Catalogue of Mental Health Measures provides a comprehensive summary of mental health and wellbeing data available from population-level longitudinal studies conducted in Britain (https://www.cataloguementalhealth.ac.uk/)

Most studies included in the reviews were large observational studies involving sample sizes in the thousands, although most reviews did not describe the study designs in detail. Those that did, described studies that were secondary analyses of existing data drawn from national or regional cohort studies, household panel surveys, repeated cross-sectional surveys, or used medical records, prescribing data or were ecological time-series studies using national or regional suicide death rates. One case-control study was included that compared mental health outcomes from a country that experienced a recession with one that did not. The reviews were, in general, not specific about which datasets were used, although one review (Martin-Carrasco et al. 2016) named two UK surveys: “The Health and Activity Lifestyle Survey” and “The English Longitudinal Study of Aging” as sources of data.

Many studies included in the reviews compared mental health in the years pre- and post-onset of an economic crisis, some described treating the Great Recession as a ‘natural experiment’. It is not clear from the reviews exactly how the date of onset of the economic crisis was determined and whether this was comparable across studies, and only two reviews (Frasquilho et al. 2016, Silva et al. 2018) gave details of specific economic measures. The macro-economic indicators that were used included national and regional rates of unemployment, gross domestic product (GDP) and home foreclosure rates.

Individual or household-level indicators included employment status, job security, household income, perceived financial stress, indebtedness, and household tenure.

The additional primary studies provided more details about study design and data sources. Of the UK-based studies, seven used data from “Understanding Society: The UK Household Longitudinal Study” (UKHLS), one used data from the “Scottish Longitudinal Study” (SLS), one from the “Welsh Health Survey” (now “National Survey for Wales”) and one from the Office for National Statistics (ONS) “Opinions and Lifestyle Survey”. The former two are panel surveys that follow the same households or individuals over time, replacing participants who leave the sample and include data linkage with external datasets, including health data. The latter two are repeated cross-sectional surveys and include a different representative sample at each wave. Economic indicators were those also identified in the reviews. In addition, primary studies in the UK measured deprivation using the Index of Multiple Deprivation, other regional income indicators and aggregated data from other household surveys such as the “Wealth and Assets Survey” and “Household Labour Survey”.

Seven of the primary studies were observational and two used a quasi-experimental design (Kim et al. 2022, Wickham et al. 2020). Both quasi-experimental studies used data from UKHLS to examine the effect of the introduction of specific welfare policies (the “Bedroom Tax” and the introduction of Universal Credit) on mental health. Wickham et al (2020) defined their “intervention group” as participants in UKHLS who answered that they were unemployed at the time Universal Credit was introduced into the area they lived in. The “control group” were defined as those who answered other than unemployed. Analysis was controlled for country of residence, age, gender, educational status, and marital status. Total sample size was 52,187. In a smaller study, but one with matched controls, Kim et al (2022), used a sub-set of participants from USS who lived in social housing in England at the time the “Bedroom Tax” was introduced and identified an “intervention group” as those living in underoccupied housing. A matched control group was identified from those not living in underoccupied housing. Total sample size was 824. Analysis was controlled for individual time-invariant variables such as immigration status and family history of immigration, and regional differences.

#### 2.2.3 Mental Health Measures

Mental health outcomes were frequently assessed using validated mental health measurement tools, which are embedded in many of the cohort and panel surveys. Twelve brief validated mental health measurement tools, self-administered, or administered by an interviewer, were identified from the included articles. These are listed in section 6 (table 4). A discussion of the psychometric properties of each tool is beyond the scope of this review, but all tools are available and widely used. The 12-item General Health Questionnaire (GHQ-12), the Short Form 12 Mental Health Component Summary (SF-12) and the Warwick-Edinburgh Mental Well-being Scale (WEMWBS) are all embedded into UKHLS, the WEMWBS is used in the “National Survey for Wales” and the 8-item Population Health Survey (PHQ-8) is used in the “Opinions and Lifestyle Survey”.

Two additional validated structured interviews were also identified, the Structured Interview to Identify Major Depressive Disorders (SCID-I) and the World Health Organisation Structured Interview to Identify Anxiety and Mood Disorder (WHO CIDI). These were used in cohort studies and are tools used to diagnose mental illness. The interviews take between 45mins to 2hrs to complete and are likely to be not practical for measuring the impact of a public health intervention on population mental health.

Additional measures of mental health included data from medical and hospital records for incidence of mental illness, prescription of medication, suicide attempts and suicide rates. Most reviews included many studies from different countries, including the UK, which used population aggregated national suicide rates in ecological time-series studies. Most reviews also included studies using data from medical records and one review (Silva et al. 2018) only included studies that examined the impact of economic crises on mental healthcare usage.

We did not try to extract data on how many studies included in the reviews used aggregated, population-level data from medical records compared to individual-level data, although it is likely that most studies used aggregated data. One grey literature report (Clark & Wenham 2022) used a combination of data from UK panel surveys, repeated cross-sectional surveys, and aggregated data from GP recorded diagnoses from the Clinical Practice Research Datalink (CRPD) and primary care data from The Health Improvement Network (THIN). We also identified one primary study that used individual-level data from SLS with linked NHS prescribing data to examine whether new anti-depressant prescriptions varied by regional economic conditions (Cherrie et al. 2021).

#### 2.2.4 Bottom line results for 2.2

Internationally a wide variety of data sources and datasets have been used to explore the impact of economic crises on mental health. Both medical records and longitudinal individual and household surveys that include validated brief mental health measurement tools, are available and widely used. The surveys aim to be representative of the population and collect information on other health-related and socio-economic outcomes. Household panel studies may include data linked to health and employment datasets, providing rich individual-level data. Within the UK, recent primary studies have tended to use the UKHLS panel survey.

### 2.3 Potential biases in study design

All reviews and studies included some discussion on strengths and limitations of study methodology (table 1).

We identified one systematic review (Saez et al. 2019) that had the aim of evaluating biases in studies assessing the effects of the Great Recession on health in Spain. This review included 53 studies and evaluated bias using an adapted tool. Four main biases were identified: problems with evaluation, time bias, failure to adjust for confounders such as seasonal effects, and inconsistencies in defining the date of onset of the Great Recession.

Problems with evaluation existed with a repeated-cross sectional design where the sample surveyed before recession onset did not consist of the same individuals as the sample surveyed after onset, and therefore were subject to confounding. Studies judged at lower risk of bias in this domain attempted to control for confounding by either matching subjects from the samples before and after the crisis; stratifying the samples by common characteristics, often into groups considered most vulnerable, or using sophisticated multilevel modelling analyses. Glonti et al (2015) also recommended other sophisticated analyses such as dynamic modelling or structural equation modelling. Other reviews also point out that it is not possible to infer causality from studies with a repeated cross-sectional design, although a strength of these studies is that they are frequently national surveys that are representative of the population. However, if a longitudinal panel survey with a repeated measures design is used, the evaluation problems do not exist, and there is stronger evidence for causality.

Time bias existed in time series studies where there were a very few data points after crisis onset, or where there was no consideration given to any potential lag between exposure and outcome. Time series studies were mainly those where the health outcome was suicide.

Other reviewers also commented that studies into suicide rates are ecological studies using aggregated data of unknown quality and that causality cannot be inferred.

Other reviewers also discussed failure or inability to adjust for confounding. A limitation of using existing datasets for secondary analysis is that data of relevance to the secondary analysis may not have been collected. For example, it was observed that UK population panel and repeated cross-sectional surveys do not always ask about both mental health and household economics in detail, and not all income- or employment-related factors of interest were included in all survey waves.

Surveys use self-reported data which may not be accurate. However, potential biases in the use of medical records were discussed in several other reviews and studies. Data in hospital and medical records were observed to be of variable quality and dependent on the structure of services and record keeping. Any reorganisation of mental health care services post-onset of an economic crisis can introduce bias by potentially altering who can access services and may disproportionate affect the most vulnerable. The use of prescribing data can introduce bias since the availability of drugs may vary, different prescribers may have different prescribing thresholds which may affect different population sub-groups differently, and the same drug may be given for conditions that are not mental-health related. One review (Silva et al. 2018) commented that people seeking medical care were not necessarily those most impacted by economic crises. Where data from medical records is aggregated, individual behaviour cannot be inferred, and the data cannot capture variation in services between population sub-groups. For example, an economic crisis may increase suicide rates for some population sub-groups but if suicide prevention strategies are provided at the same time, these may benefit different population sub-groups and the net result may be a zero increase. However, it was observed that an advantage of aggregated data was that they reflect the environmental effects of changes in the economy beyond an individual’s personal circumstances.

#### 2.3.1 Bottom line results for 2.3

There are strengths and weaknesses and the potential to introduce bias in all study designs. The choice of study design is likely to be a pragmatic decision based on the exact research question and availability of data. The use of data drawn from large population surveys, especially when linked to medical records is efficient and can ensure that the sample is representative of the population. However, care should be taken if data on important confounders is missing.

### 2.4 Vulnerable groups

All the reviews and grey literature and some primary studies identified population groups whose mental health was most likely to be affected by an economic crisis. All the mental health measurement tools reported in section 2.2 were used for this purpose, except for the WEMWBS. Tables 3a to 3c summarises the vulnerable groups identified in each review and primary study (section 6). All the reviews presented the data narratively and neither reviews nor primary studies commented on the suitability of the methods and tools when identifying vulnerable groups. However, consistently, the most vulnerable groups were identified as those who were living on lower incomes, in financial or housing insecurity or living in more deprived areas prior to the crisis. People who were unemployed or whose employment was precarious were at high risk of worsening mental health problems.

The reviews also examined data stratified by gender and age. It appeared that men’s mental health deteriorated more during economic crises, but women were more likely to experience poorer mental health at all timepoints and were more likely to access mental health services. However, men, particularly men of working age were at higher risk of suicide. Some studies linked changes in mental health in men and women to socio-economic status but not all and the evidence for age-related differences was mixed. Other groups identified as being at risk were people with pre-existing mental health problems, people living in households with dependent children, and migrants. One primary study (Office for National Statistics 2022) also identified adults with a disability or long-term illness as being at risk. No included study or review presented data from people from minority ethnic groups.

It appears that the methods and tools are suitable for use in men and women, across the socio-economic spectrum, living in different housing tenures and with different employment status. However, it is possible that some potentially vulnerable groups are not well represented when using the methods and data sources identified.

## 3. DISCUSSION

### 3.1 Summary of the findings

This rapid review aimed to answer two questions:

1. **What methods and tools are available and appropriate for monitoring the impact of the cost-of-living crisis on mental health?**

We identified multiple studies that examined the impact of economic crises on mental health over time using national surveys from many different countries using different methods. Data from population-level panel surveys may be most useful. The UK has many ongoing population-level surveys, providing rich data that can be used to explore the health of the population over time. These large surveys incorporate validated mental health tools and questions to determine financial security at household or individual level and may be linked to medical records and CPRD data. Several primary studies published since 2021 used data from UKHLS. Many studies are observational in design, but we also identified two relevant quasi-experimental case-control studies, that used UKHLS data to examine the impact of specific welfare policies in the UK (the “Bedroom tax” and the introduction of Universal Credit). A quasi-experimental study design, using UKHLS data, could also be suitable for measuring the impact of a specific public health initiative that has a clear roll-out date.

2. **Are methods and mental health measurement tools suitable for specific, vulnerable populations?**

All the reviews and most primary studies found people living in financial insecurity were at higher risk of poor or worsening mental health and most examined gender differences. It therefore appears that the methods and tools used were suitable for capturing data from men and women, from different socio-economic groups. Few studies presented data from other groups that are often marginalised such as migrants and people living with disabilities or long-term poor health. We also identified no studies that examined whether people from minority ethnic groups were more at risk of deteriorating mental health during economic crises. It is therefore not clear as to whether the methods identified are suitable for all marginalised populations.

### 3.2 Strengths and limitations of this Rapid Review

Evidence for this rapid review has been drawn from reviews that included over 300 studies, and from primary studies and grey literature that were of relevance to Wales. We have identified appropriate available population level datasets that include both validated mental health tools and collect socioeconomic outcomes and have identified appropriate study designs and provided information about their strengths and limitations.

However, our rapid review has limitations. All the included reviews summarised data from individual studies narratively and varied in the detail they provided on study designs and strengths and limitations. The identification of vulnerable groups appeared not to be systematic in most reviews and it is possible that data on vulnerable groups was not captured. There is a strong overlap in the literature with studies describing the impact of economic crises on mental health and the impact of any austerity policies that followed the Great Recession on mental health. We did not include austerity as a search term, and it is possible that relevant studies were excluded. However, the purpose of this review was not to examine the evidence that either economic crises or austerity policies affect mental health, but to describe available methods of measuring changes to population mental health in response to public health intervention in Wales. In addition, although the search strategy was comprehensive and included evidence from peer reviewed articles and from grey literature, we had to exclude much of the identified grey literature due to lack of methodological detail and lack of information on data sources and measurement tools. It is possible that there are datasets and tools used by charities and institutions that we did not identify.

### 3.3 Implications for policy and research

Our rapid review has identified existing methods and tools likely to be suitable for measuring the impact of public health initiatives on mental health in people from different socio-economic groups during a cost-of-living crisis. However, it is unclear as to whether the identified methods and tools adequately capture data from people from minority ethnic groups, who already experience disparities in mental health care (Ahmad et al. 2022). In the short-term, reports and studies using panel surveys such as UKHLS or that use medical records should present data on ethnicity and, where possible, plan to stratify analyses by ethnicity. In the longer-term it may be necessary to develop new methods to better capture data from people from minority ethnic groups.

### 3.4 Economic considerations^*^

- The UK has seen sharp rises in inflation since Winter 2021. The Consumer Price Index including owner occupier housing costs (CPIH) rose from 3% in August 2021 to a peak 9.6% in October 2022. CPIH is currently 7.9% (Office for National Statistics, 2023).
- In the lead up to the cost of living crisis, Wales had the highest proportion of working age adults (21%) and pensioners (18%) in relative income poverty out of the UK nations. 28% of children in Wales were living in relative poverty (Welsh Government, 2023a). Given that over half of all mental health problems start by age 14 (and 75% by age 18) and poverty being a known risk factor for psychological illnesses, there is likely to be a long shadow of mental health continuing into future generations stemming from the cost of living crisis (Department of Health, 2015; Fell & Hewstone, 2015).
- The effects of the cost of living crisis are not felt equally. Poorer households tend to spend a greater proportion of income on items more exposed to inflationary pressures such as energy and food (Senedd Research, 2022). 90% of people in Wales felt their health was negatively affected due to increased heating costs (Royal College of Physicians, 2022). Decisions around the use of energy at home can lead to lingering anxiety that negatively impacts mental health (Marmot, 2020).
- Mental Health problems cost the Welsh economy £4.8 billion per annum. 72% of these incurred costs are attributed to productivity losses of people living with mental health conditions and costs experienced by unpaid informal carers (Mcdaid et al., 2022).
- In a survey of 2,000 Welsh participants covering the period November 2022 to January 2023, 60% of respondents agreed that rising costs of living negatively affected their quality of life (25% strongly agreed). 87% reported ‘worrying’ around the cost of living, with 38% reporting ‘worrying a lot’ (Public Health Wales, 2023).
- People with poor mental health are more likely to experience subsequent reductions in income. The negative impacts on mental health and well-being induced by the cost of living crisis could lead to future financial and health problems. Potentially creating a downward spiral even that can persist even if economic conditions improve (Thomson et al., 2022).
- Practitioners in Wales have voiced concerns over the compounding socioeconomic consequences of the COVID-19 pandemic and the subsequent cost of living crisis affecting the mental health of the Welsh population (Welsh Government, 2023b).

## Data Availability

All data produced in the present study are available upon reasonable request to the authors

## Abbreviations

CRPD: Clinical Practice Research Datalink
GHQ-12: 12-item General Health Questionnaire
GDP: Gross Domestic Product
ONS: Office for National Statistics
PHQ-8: Population Health Survey
SF-12: Short Form 12 Mental Health Component Summary
SLS: Scottish Longitudinal Study
THIN: The Health Improvement Network
UKHLS: Understanding Society: The UK Household Longitudinal Study
WEMWBS: Warwick-Edinburgh Mental Well-being Scale

## 5. RAPID REVIEW METHODS

### 5.1 Eligibility criteria

**Table 2:** Eligibility criteria.

|  | **Inclusion criteria** | **Exclusion criteria** |
| --- | --- | --- |
| **Population** | Adults or households | Organisations and systems (e.g., exploring how an organisation such as a health / educational organisation or system responds)<br>Children/adolescents only |
| **Exposure** | Cost-of-living crisis or economic / financial crisis | Related to financial stress caused by Covid-19 and/or measures to tackle Covid-19 or other infectious disease or disaster (natural or man-made) |
| **Outcome** | Mental health e.g.,<br>• Depressive/affective disorders<br>• Anxiety disorders<br>• Disruptive behaviour and dissocial disorders<br>• Stress<br>• Wellbeing<br>• Suicide attempts/ideation<br>• Suicide completion rates | Physical health<br>Behavioural outcomes (such as healthy eating, physical activity)<br>Sleep outcomes<br>Psychosis<br>Schizophrenia<br>Substance misuse<br>Eating disorders |
| **Evaluation** | Appraisal of the following:<br>• Validated population-level surveys<br>• Population-level mental healthcare use (including, but not limited to primary and secondary care records, hospital admissions, medication use) | Appraisal of questionnaires used only for individual level data.<br>Non-validated tools developed specifically for a study (bespoke tools) |
| **Study design** | • Systematic reviews; scoping reviews; rapid reviews, organisational reports; clinical guidance<br>Primary research, limited to:<br>• Longitudinal observational studies (panel surveys, time series, interrupted time series and qualitative studies)<br>• Repeated, population-level cross-sectional studies<br>• Population-level intervention studies (randomised, non-randomised and single arm) | Single timepoint cross-sectional studies<br>Individual-level intervention studies |
| **Countries** | Conducted in UK, Europe, Australia, New Zealand. | All other countries |
| **Language of publication** | English |  |
| **Publication date** | Secondary evidence from 1970<br>Primary studies 2021- present |  |
| **Publication type** | Published, including grey literature | Conference abstracts<br>Preprints |
| **Other factors** | Prior to 2021 relevant measures will be identified from secondary evidence (systematic reviews; scoping reviews; rapid reviews; rapid evidence assessments; organisational reports; clinical guidance). This date limit was arrived at since preparatory work had identified a large, eligible, scoping review that included articles published up to the end of Dec 2020 |  |

### 5.2 Literature search

Studies that evaluated the impact of economic crises on mental health were identified through electronic searches conducted between 27^th^ March 2023 and 6^th^ April 2023 in the following databases: KSR Evidence; Ovid Medline, Embase, PsycInfo, SSP and HMIC; EbscoHOST CINAHL; Scopus; Wiley Cochrane; ProQuest ASSIA, SSA, SA and SD; Social Care Online; TripPro; PROSPERO; Google and Google Scholar. The searches strategy was developed by two information specialists, with input from the lead author and carried out in two parts:

1. Cost-of-living & mental health (main evidence search)

a. secondary evidence (1970–2023) – standard HTW systematic review/meta-analysis & guidelines/HTA filters were be applied
b. primary evidence (2021-2023) – the systematic review set 1a were excluded from these results, no other filters were applied
2. Cost of living mop-up searches

a. cost of living and psychological assessment tools (1970-2023)
b. cost of living and Wales (1970-2023)

Full details of the searches and search terms can be found in the appendix.

### 5.3 Study selection process

All references identified by the literature searches were screened for eligibility using the criteria above by title and abstract. Where it was deemed that the title and abstract was potentially relevant, the full text of articles was obtained and screened.

Firstly, existing reviews describing the effect of the cost-of-living crisis on mental health were assessed for relevance using their inclusion/exclusion criteria and characteristics of included studies. Reviews that did not clearly define their inclusion/exclusion criteria or where the majority of included studies did not fit our eligibility criteria (e.g., not conducted in the countries of interest or mainly cross-sectional studies) or that did not include a description of measurement tools were excluded. Whilst reviews that evaluated methods of measuring mental health at a population level or their responsiveness to interventions were considered for inclusion in the synthesis. The inclusion of reviews with a minority of studies that did not meet our inclusion criteria was a pragmatic decision because we were aware that many individual studies in this field were cross-sectional, and few reviews existed that only included longitudinal studies.

Due to the number of relevant articles identified, it was not possible to include all reviews and studies. Where multiple high-quality reviews were found that could answer the research questions, the largest or most recent reviews that included details of the available tools and methods were chosen for inclusion. Relevant full texts of primary studies from 2021 to April 2023 were screened and included according to the following criteria 1) used evaluation methods not previously identified from reviews 2) with an experimental/quasi-experimental design 3) studies of relevance to Wales 4) included detailed exploration of strengths and limitations of evaluation methods. These decisions meant that most of the included primary studies and all the grey literature were UK-based.

Screening was conducted by a single reviewer. A second reviewer quality assured 20% of the primary studies.

### 5.4 Data extraction

The following data were extracted:

- Study details (author, year, country, purpose, design)
- Secondary evidence: Number of included studies and key characteristics
- Mental health measurement methods and tools
- Economic measures
- Sources of data
- Strengths and limitations of the measurement tools as identified by study authors
- Vulnerable groups as identified by study authors

Where existing reviews were used, data specific to the measurement tools and other evaluation methods was extracted either directly from the review or from included primary studies that met our inclusion criteria where the review lacks all relevant detail. Extracted data included any evaluation of the measurement tools conducted by review authors (table 3).

Where necessary further, non-systematic, searches were conducted to obtain details of the characteristics of the identified measurement tools and methods, prioritising articles describing the original development of the tool. A full review and quality assessment of the psychometric properties of each tool was beyond the scope of this rapid review, instead a brief description of each tool, including domain, number of items, time taken to administration and availability was provided (table 4)

Data were extracted by a single reviewer, with 20% of records quality assured by a second reviewer.

### 5.5 Quality appraisal

The purpose of this rapid review was to identify available methods and tools for evaluating the impact of the cost-of-living crisis on mental health. Formal quality appraisal of the evidence presented by the reviews and studies that used the methods was beyond the scope of the rapid review. A narrative description of the search strategy and inclusion criteria of included evidence is provided in table 3 and discussion of the potential study designs using the methods identified (table 1).

### 5.6 Synthesis

A narrative synthesis on relevant methods, the context in which they are used, and the strengths and limitations as identified by study authors was presented.

## 6. EVIDENCE

### 6.1 Search results and study selection

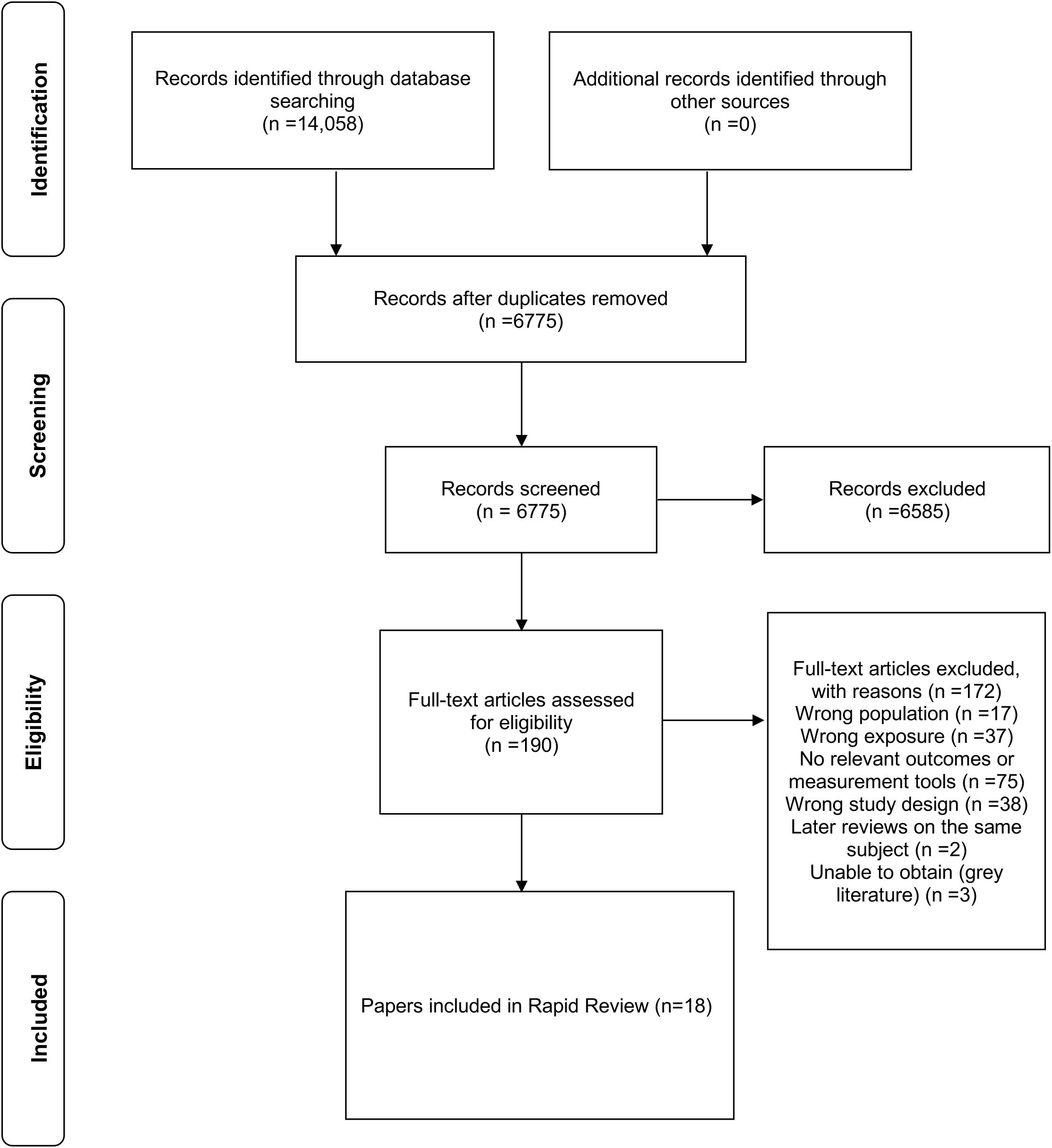

### 6.2 Data extraction

Tables 3a to 3c present the data extracted from the included reviews and studies. Table 4 presents an overview of identified validated mental health measurement tools.

#### 6.2.1 Summary of evidence

**Table 3a.**
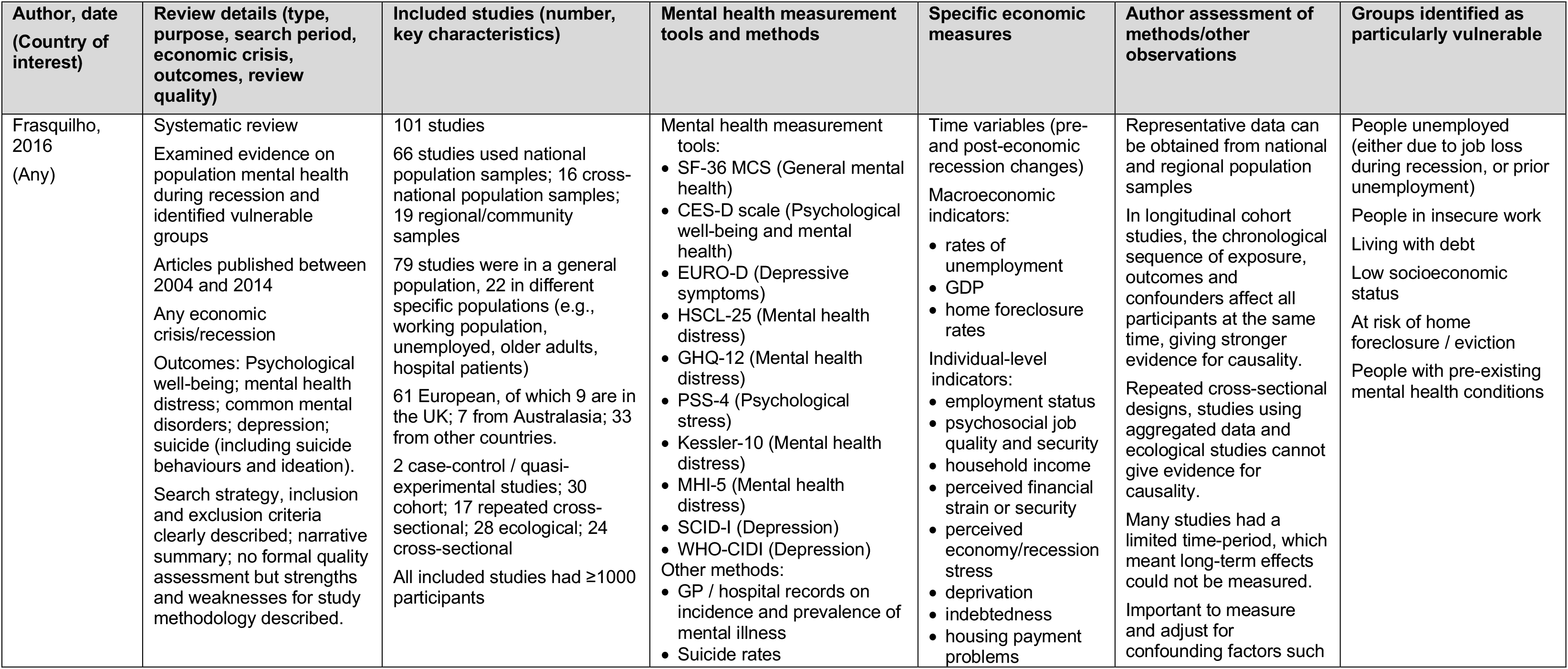
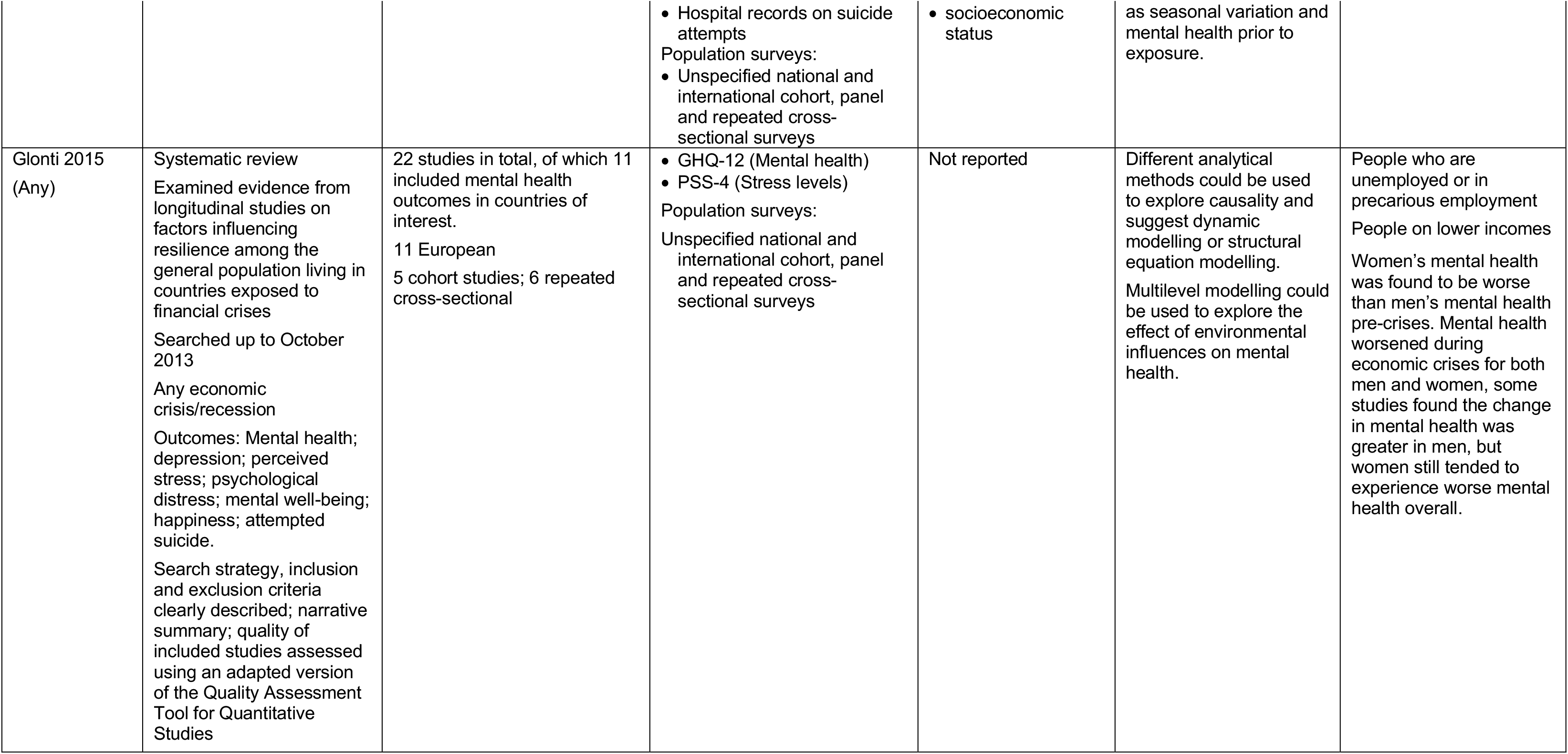
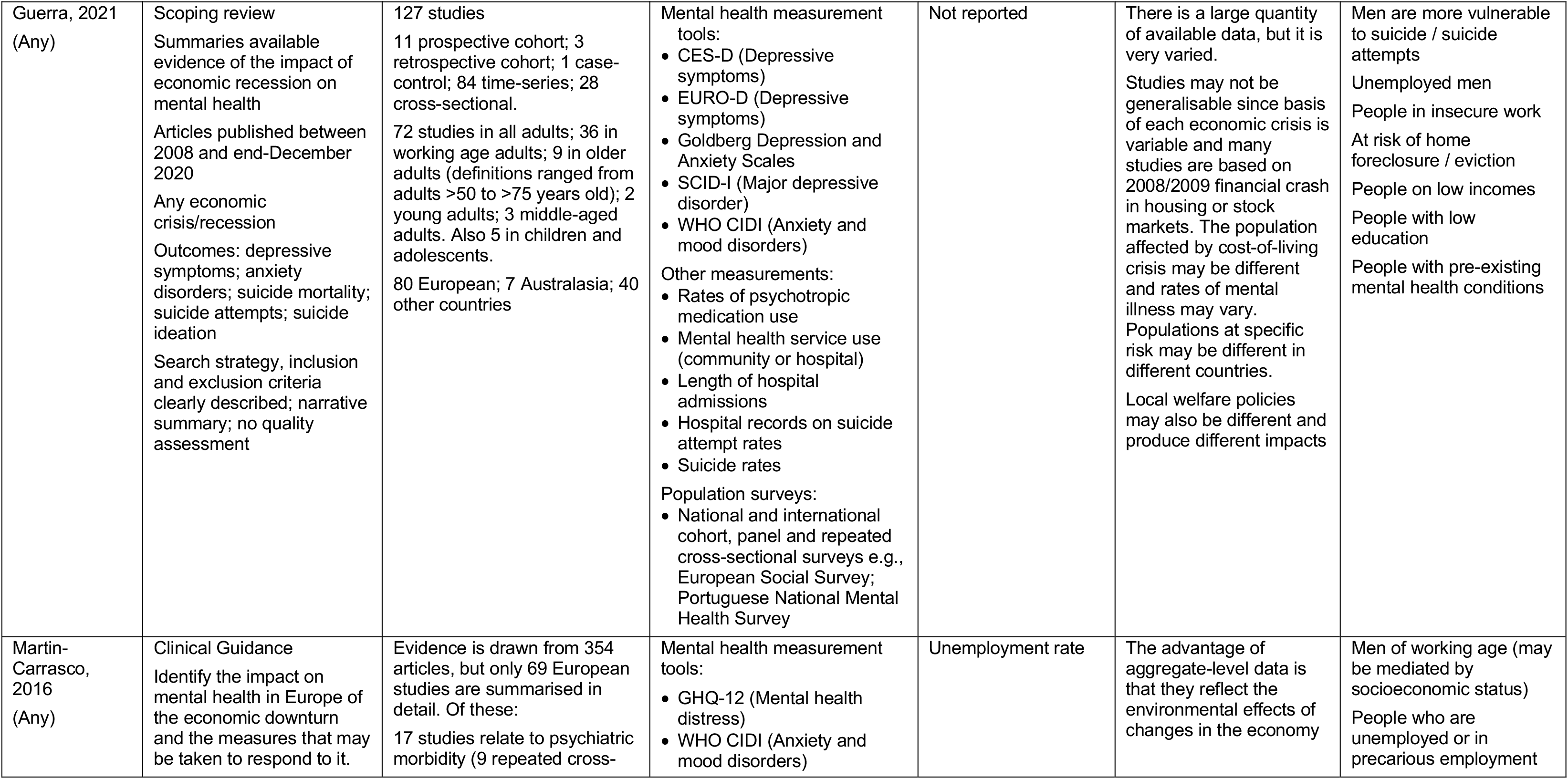
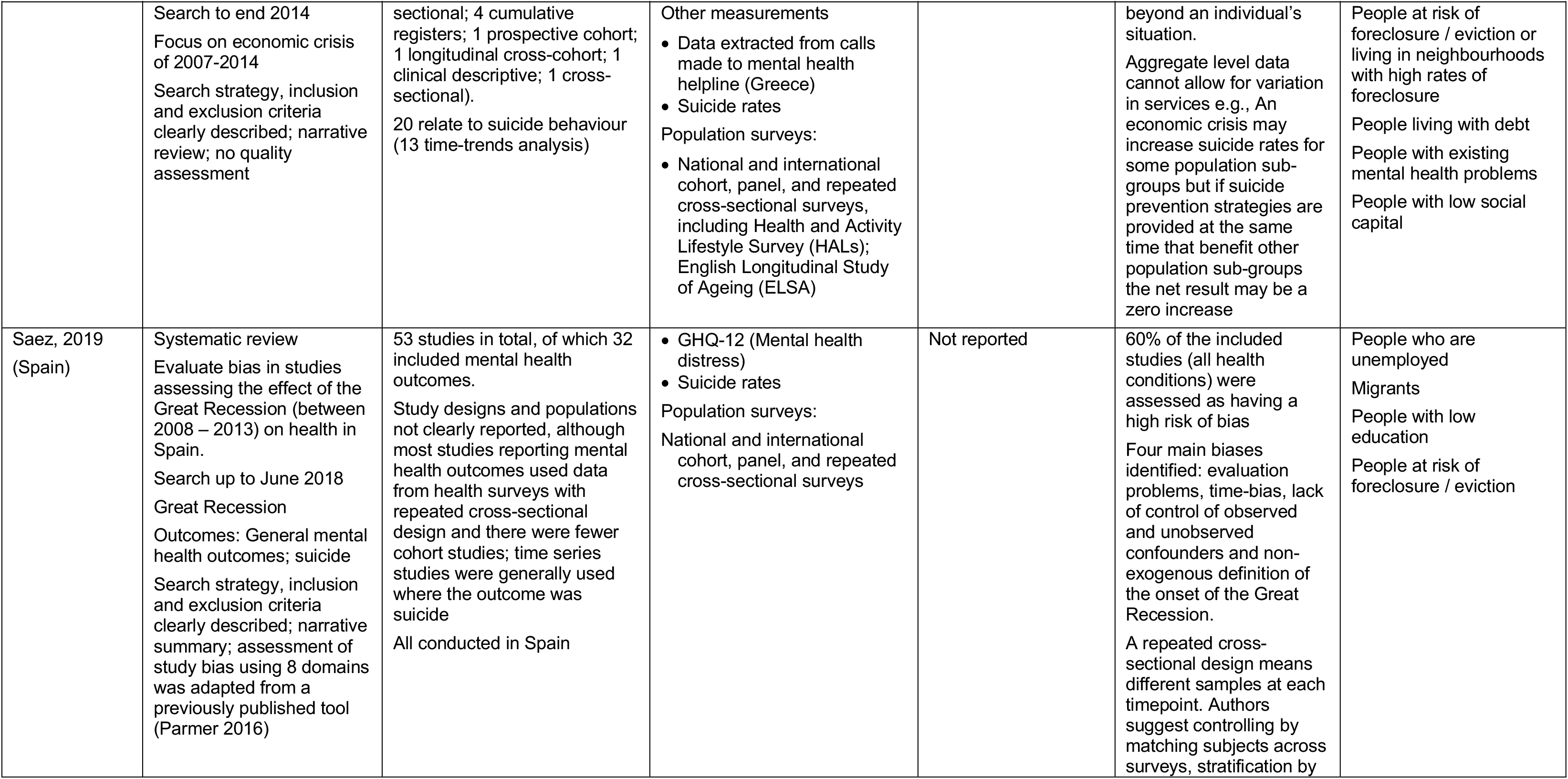
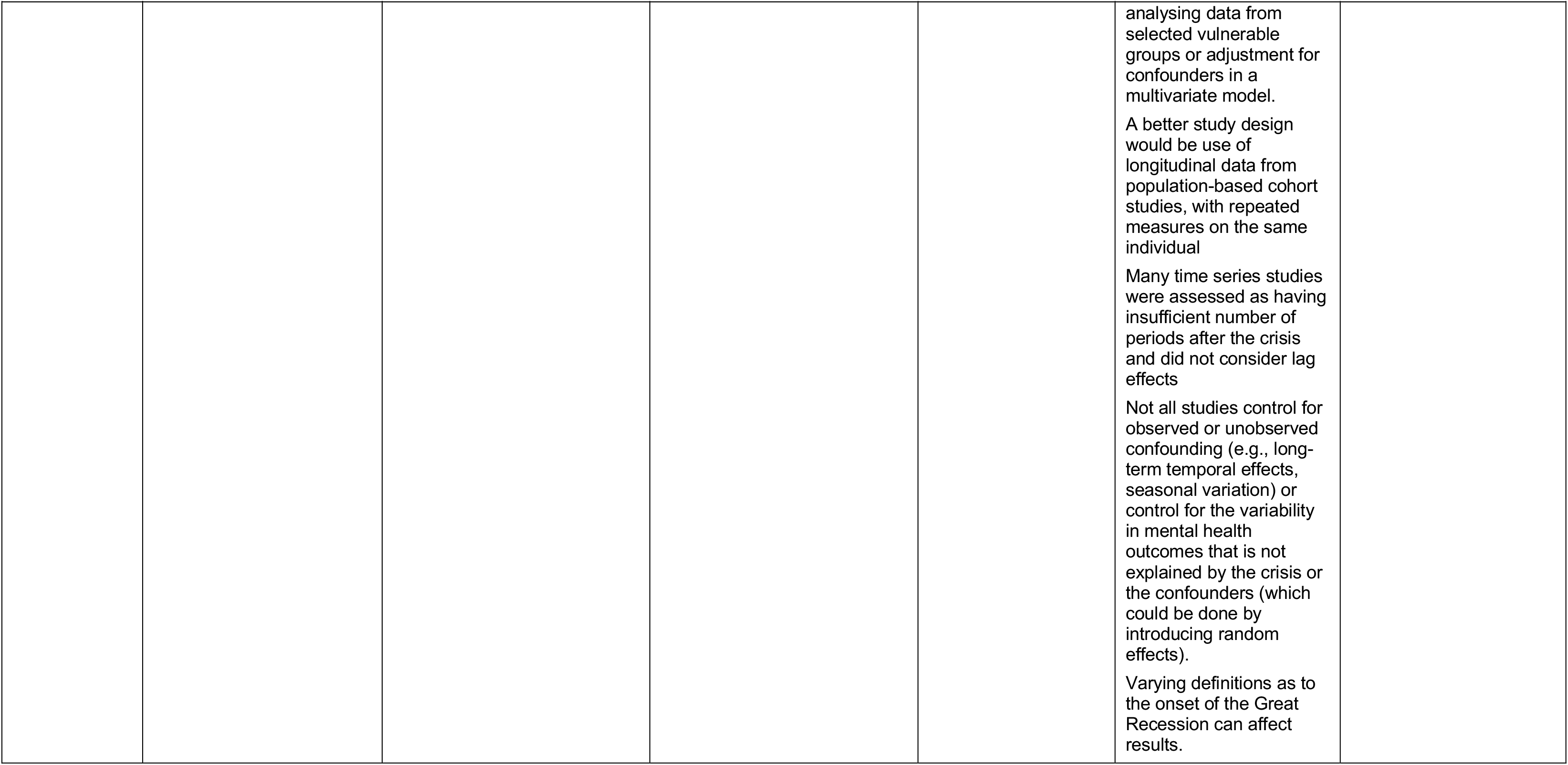
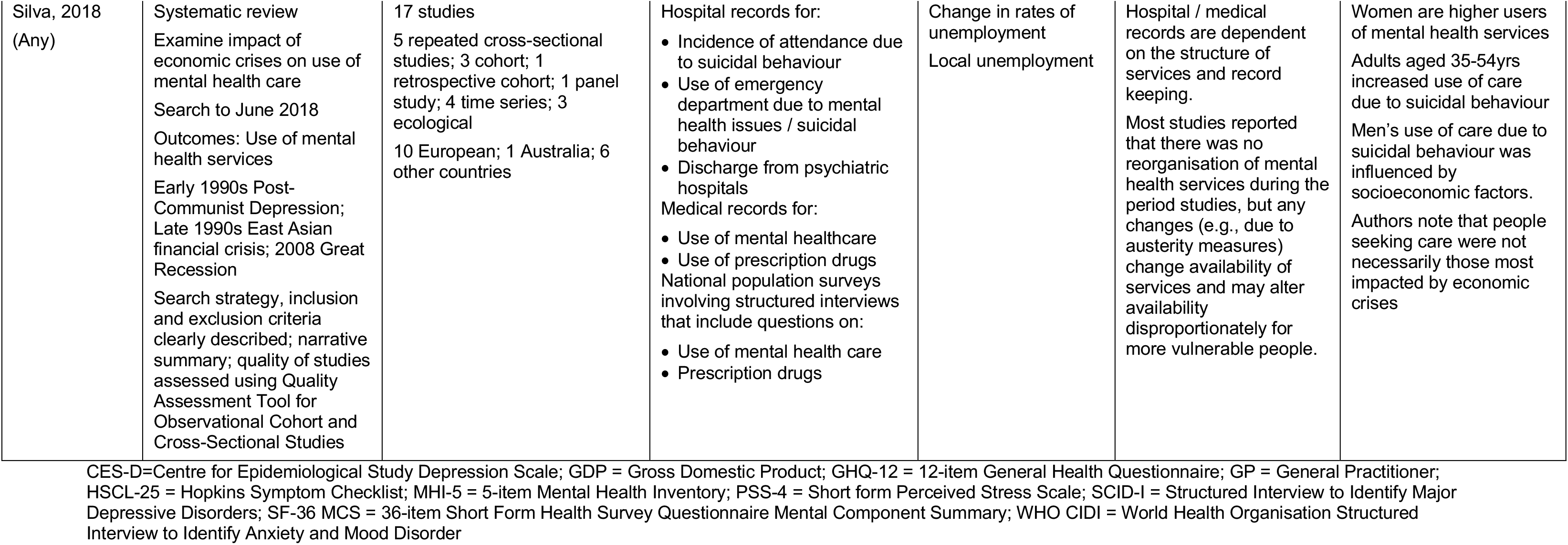
Peer-reviewed secondary evidence.

| Author, date<br>(Country of interest) | Review details (type, purpose, search period, economic crisis, outcomes, review quality) | Included studies (number, key characteristics) | Mental health measurement tools and methods | Specific economic measures | Author assessment of methods/other observations | Groups identified as particularly vulnerable |
| --- | --- | --- | --- | --- | --- | --- |
| Frasquilho, 2016<br>(Any) | Systematic review<br>Examined evidence on population mental health during recession and identified vulnerable groups<br>Articles published between 2004 and 2014<br>Any economic crisis/recession<br>Outcomes: Psychological well-being; mental health distress; common mental disorders; depression; suicide (including suicide behaviours and ideation).<br>Search strategy, inclusion and exclusion criteria clearly described; narrative summary; no formal quality assessment but strengths and weaknesses for study methodology described. | 101 studies<br>66 studies used national population samples; 16 cross-national population samples; 19 regional/community samples<br>79 studies were in a general population, 22 in different specific populations (e.g., working population, unemployed, older adults, hospital patients)<br>61 European, of which 9 are in the UK; 7 from Australasia; 33 from other countries.<br>2 case-control / quasi-experimental studies; 30 cohort; 17 repeated cross-sectional; 28 ecological; 24 cross-sectional<br>All included studies had ≥1000 participants | Mental health measurement tools:<br>• SF-36 MCS (General mental health)<br>• CES-D scale (Psychological well-being and mental health)<br>• EURO-D (Depressive symptoms)<br>• HSCL-25 (Mental health distress)<br>• GHQ-12 (Mental health distress)<br>• PSS-4 (Psychological stress)<br>• Kessler-10 (Mental health distress)<br>• MHI-5 (Mental health distress)<br>• SCID-I (Depression)<br>• WHO-CIDI (Depression)<br>Other methods:<br>• GP / hospital records on incidence and prevalence of mental illness<br>• Suicide rates | Time variables (pre- and post-economic recession changes)<br>Macroeconomic indicators:<br>• rates of unemployment<br>• GDP<br>• home foreclosure rates<br>Individual-level indicators:<br>• employment status<br>• psychosocial job quality and security<br>• household income<br>• perceived financial strain or security<br>• perceived economy/recession stress<br>• deprivation<br>• indebtedness<br>• housing payment problems | Representative data can be obtained from national and regional population samples<br>In longitudinal cohort studies, the chronological sequence of exposure, outcomes and confounders affect all participants at the same time, giving stronger evidence for causality.<br>Repeated cross-sectional designs, studies using aggregated data and ecological studies cannot give evidence for causality.<br>Many studies had a limited time-period, which meant long-term effects could not be measured.<br>Important to measure and adjust for confounding factors such | People unemployed (either due to job loss during recession, or prior unemployment)<br>People in insecure work<br>Living with debt<br>Low socioeconomic status<br>At risk of home foreclosure / eviction<br>People with pre-existing mental health conditions |
|  |  |  | Hospital records on suicide attempts<br>Population surveys:<br>Unspecified national and international cohort, panel and repeated cross-sectional surveys | socioeconomic status | as seasonal variation and mental health prior to exposure. |  |
| Glonti 2015<br>(Any) | Systematic review<br>Examined evidence from longitudinal studies on factors influencing resilience among the general population living in countries exposed to financial crises<br>Searched up to October 2013<br>Any economic crisis/recession<br>Outcomes: Mental health; depression; perceived stress; psychological distress; mental well-being; happiness; attempted suicide.<br>Search strategy, inclusion and exclusion criteria clearly described; narrative summary; quality of included studies assessed using an adapted version of the Quality Assessment Tool for Quantitative Studies | 22 studies in total, of which 11 included mental health outcomes in countries of interest.<br>11 European<br>5 cohort studies; 6 repeated cross-sectional | GHQ-12 (Mental health)<br>PSS-4 (Stress levels)<br>Population surveys:<br>Unspecified national and international cohort, panel and repeated cross-sectional surveys | Not reported | Different analytical methods could be used to explore causality and suggest dynamic modelling or structural equation modelling.<br>Multilevel modelling could be used to explore the effect of environmental influences on mental health. | People who are unemployed or in precarious employment<br>People on lower incomes<br>Women's mental health was found to be worse than men's mental health pre-crises. Mental health worsened during economic crises for both men and women, some studies found the change in mental health was greater in men, but women still tended to experience worse mental health overall. |
| Guerra, 2021<br>(Any) | Scoping review<br>Summaries available evidence of the impact of economic recession on mental health<br>Articles published between 2008 and end-December 2020<br>Any economic crisis/recession<br>Outcomes: depressive symptoms; anxiety disorders; suicide mortality; suicide attempts; suicide ideation<br>Search strategy, inclusion and exclusion criteria clearly described; narrative summary; no quality assessment | 127 studies<br>11 prospective cohort; 3 retrospective cohort; 1 case-control; 84 time-series; 28 cross-sectional.<br>72 studies in all adults; 36 in working age adults; 9 in older adults (definitions ranged from adults >50 to >75 years old); 2 young adults; 3 middle-aged adults. Also 5 in children and adolescents.<br>80 European; 7 Australasia; 40 other countries | Mental health measurement tools:<br>• CES-D (Depressive symptoms)<br>• EURO-D (Depressive symptoms)<br>• Goldberg Depression and Anxiety Scales<br>• SCID-I (Major depressive disorder)<br>• WHO CIDI (Anxiety and mood disorders)<br>Other measurements:<br>• Rates of psychotropic medication use<br>• Mental health service use (community or hospital)<br>• Length of hospital admissions<br>• Hospital records on suicide attempt rates<br>• Suicide rates<br>Population surveys:<br>• National and international cohort, panel and repeated cross-sectional surveys e.g., European Social Survey; Portuguese National Mental Health Survey | Not reported | There is a large quantity of available data, but it is very varied.<br>Studies may not be generalisable since basis of each economic crisis is variable and many studies are based on 2008/2009 financial crash in housing or stock markets. The population affected by cost-of-living crisis may be different and rates of mental illness may vary.<br>Populations at specific risk may be different in different countries.<br>Local welfare policies may also be different and produce different impacts | Men are more vulnerable to suicide / suicide attempts<br>Unemployed men<br>People in insecure work<br>At risk of home foreclosure / eviction<br>People on low incomes<br>People with low education<br>People with pre-existing mental health conditions |
| Martin-Carrasco, 2016<br>(Any) | Clinical Guidance<br>Identify the impact on mental health in Europe of the economic downturn and the measures that may be taken to respond to it. | Evidence is drawn from 354 articles, but only 69 European studies are summarised in detail. Of these:<br>17 studies relate to psychiatric morbidity (9 repeated cross- | Mental health measurement tools:<br>• GHQ-12 (Mental health distress)<br>• WHO CIDI (Anxiety and mood disorders) | Unemployment rate | The advantage of aggregate-level data is that they reflect the environmental effects of changes in the economy | Men of working age (may be mediated by socioeconomic status)<br>People who are unemployed or in precarious employment |
|  | Search to end 2014<br>Focus on economic crisis of 2007-2014<br>Search strategy, inclusion and exclusion criteria clearly described; narrative review; no quality assessment | sectional; 4 cumulative registers; 1 prospective cohort; 1 longitudinal cross-cohort; 1 clinical descriptive; 1 cross-sectional).<br>20 relate to suicide behaviour (13 time-trends analysis) | Other measurements<br>• Data extracted from calls made to mental health helpline (Greece)<br>• Suicide rates<br>Population surveys:<br>• National and international cohort, panel, and repeated cross-sectional surveys, including Health and Activity Lifestyle Survey (HALs); English Longitudinal Study of Ageing (ELSA) |  | beyond an individual's situation.<br>Aggregate level data cannot allow for variation in services e.g., An economic crisis may increase suicide rates for some population sub-groups but if suicide prevention strategies are provided at the same time that benefit other population sub-groups the net result may be a zero increase | People at risk of foreclosure / eviction or living in neighbourhoods with high rates of foreclosure<br>People living with debt<br>People with existing mental health problems<br>People with low social capital |
| Saez, 2019<br>(Spain) | Systematic review<br>Evaluate bias in studies assessing the effect of the Great Recession (between 2008 – 2013) on health in Spain.<br>Search up to June 2018<br>Great Recession<br>Outcomes: General mental health outcomes; suicide<br>Search strategy, inclusion and exclusion criteria clearly described; narrative summary; assessment of study bias using 8 domains was adapted from a previously published tool (Parmer 2016) | 53 studies in total, of which 32 included mental health outcomes.<br>Study designs and populations not clearly reported, although most studies reporting mental health outcomes used data from health surveys with repeated cross-sectional design and there were fewer cohort studies; time series studies were generally used where the outcome was suicide<br>All conducted in Spain | • GHQ-12 (Mental health distress)<br>• Suicide rates<br>Population surveys: National and international cohort, panel, and repeated cross-sectional surveys | Not reported | 60% of the included studies (all health conditions) were assessed as having a high risk of bias<br>Four main biases identified: evaluation problems, time-bias, lack of control of observed and unobserved confounders and non-exogenous definition of the onset of the Great Recession.<br>A repeated cross-sectional design means different samples at each timepoint. Authors suggest controlling by matching subjects across surveys, stratification by<br>analysing data from selected vulnerable groups or adjustment for confounders in a multivariate model.<br>A better study design would be use of longitudinal data from population-based cohort studies, with repeated measures on the same individual<br>Many time series studies were assessed as having insufficient number of periods after the crisis and did not consider lag effects<br>Not all studies control for observed or unobserved confounding (e.g., long-term temporal effects, seasonal variation) or control for the variability in mental health outcomes that is not explained by the crisis or the confounders (which could be done by introducing random effects).<br>Varying definitions as to the onset of the Great Recession can affect results. | People who are unemployed<br>Migrants<br>People with low education<br>People at risk of foreclosure / eviction |
| Silva, 2018<br>(Any) | Systematic review<br>Examine impact of economic crises on use of mental health care<br>Search to June 2018<br>Outcomes: Use of mental health services<br>Early 1990s Post-Communist Depression; Late 1990s East Asian financial crisis; 2008 Great Recession<br>Search strategy, inclusion and exclusion criteria clearly described; narrative summary; quality of studies assessed using Quality Assessment Tool for Observational Cohort and Cross-Sectional Studies | 17 studies<br>5 repeated cross-sectional studies; 3 cohort; 1 retrospective cohort; 1 panel study; 4 time series; 3 ecological<br>10 European; 1 Australia; 6 other countries | Hospital records for:<br>Incidence of attendance due to suicidal behaviour<br>Use of emergency department due to mental health issues / suicidal behaviour<br>Discharge from psychiatric hospitals<br>Medical records for:<br>Use of mental healthcare<br>Use of prescription drugs<br>National population surveys involving structured interviews that include questions on:<br>Use of mental health care<br>Prescription drugs | Change in rates of unemployment<br>Local unemployment | Hospital / medical records are dependent on the structure of services and record keeping.<br>Most studies reported that there was no reorganisation of mental health services during the period studies, but any changes (e.g., due to austerity measures) change availability of services and may alter availability disproportionately for more vulnerable people. | Women are higher users of mental health services<br>Adults aged 35-54yrs increased use of care due to suicidal behaviour<br>Men's use of care due to suicidal behaviour was influenced by socioeconomic factors.<br>Authors note that people seeking care were not necessarily those most impacted by economic crises |

CES-D=Centre for Epidemiological Study Depression Scale; GDP = Gross Domestic Product; GHQ-12 = 12-item General Health Questionnaire; GP = General Practitioner; HSCL-25 = Hopkins Symptom Checklist; MHI-5 = 5-item Mental Health Inventory; PSS-4 = Short form Perceived Stress Scale; SCID-I = Structured Interview to Identify Major Depressive Disorders; SF-36 MCS = 36-item Short Form Health Survey Questionnaire Mental Component Summary; WHO CIDI = World Health Organisation Structured Interview to Identify Anxiety and Mood Disorder

**Table 3b.**
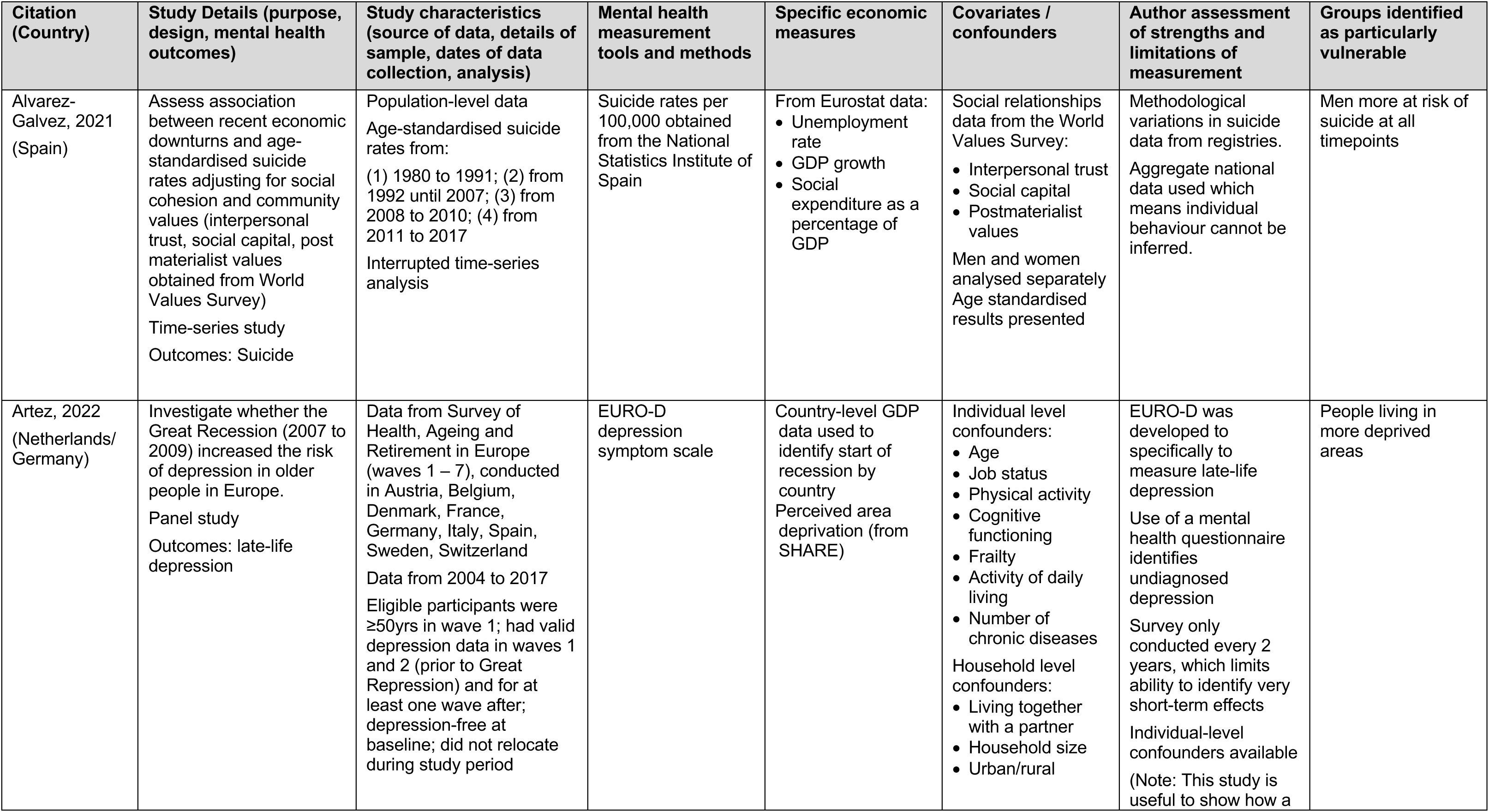
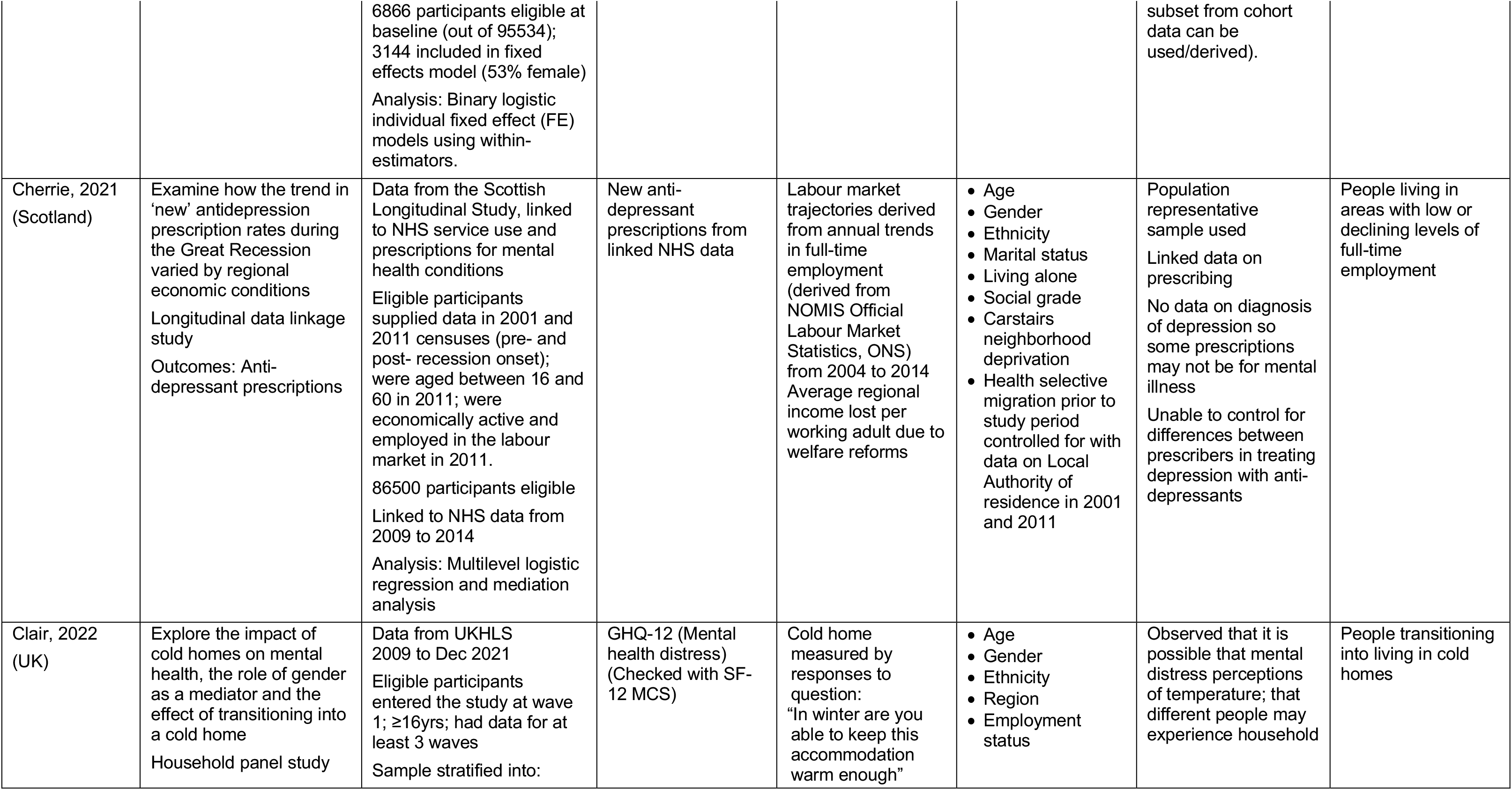
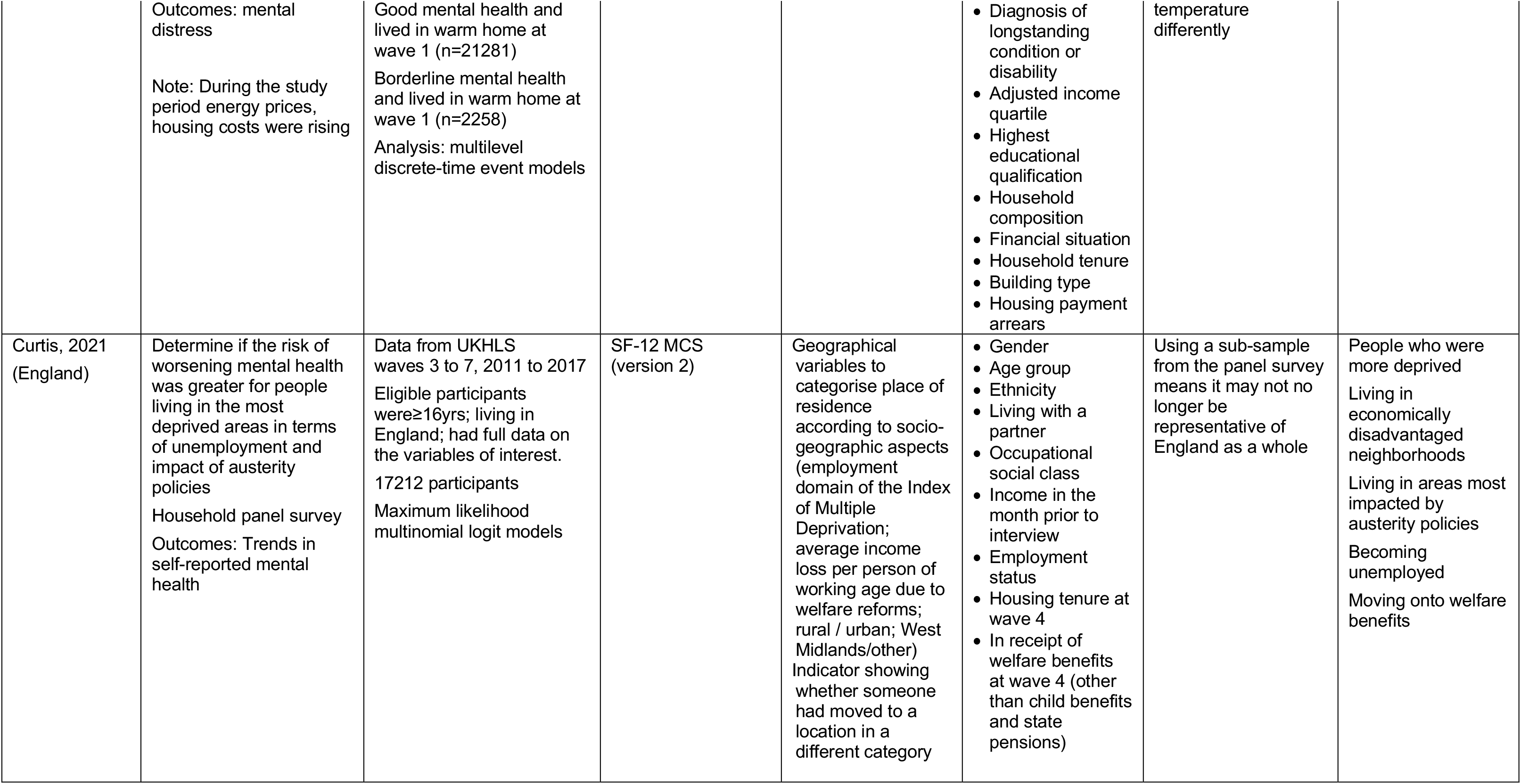
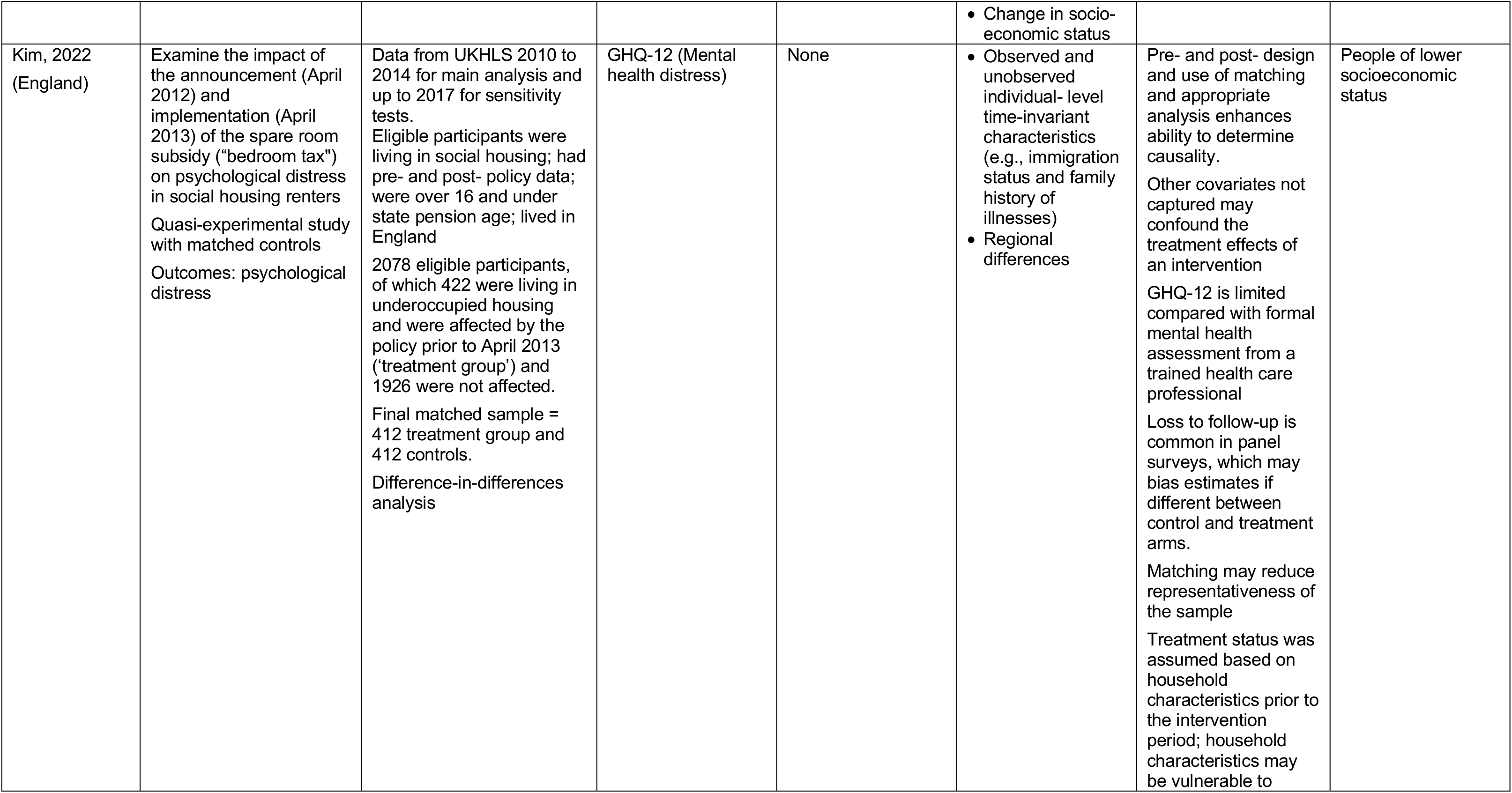
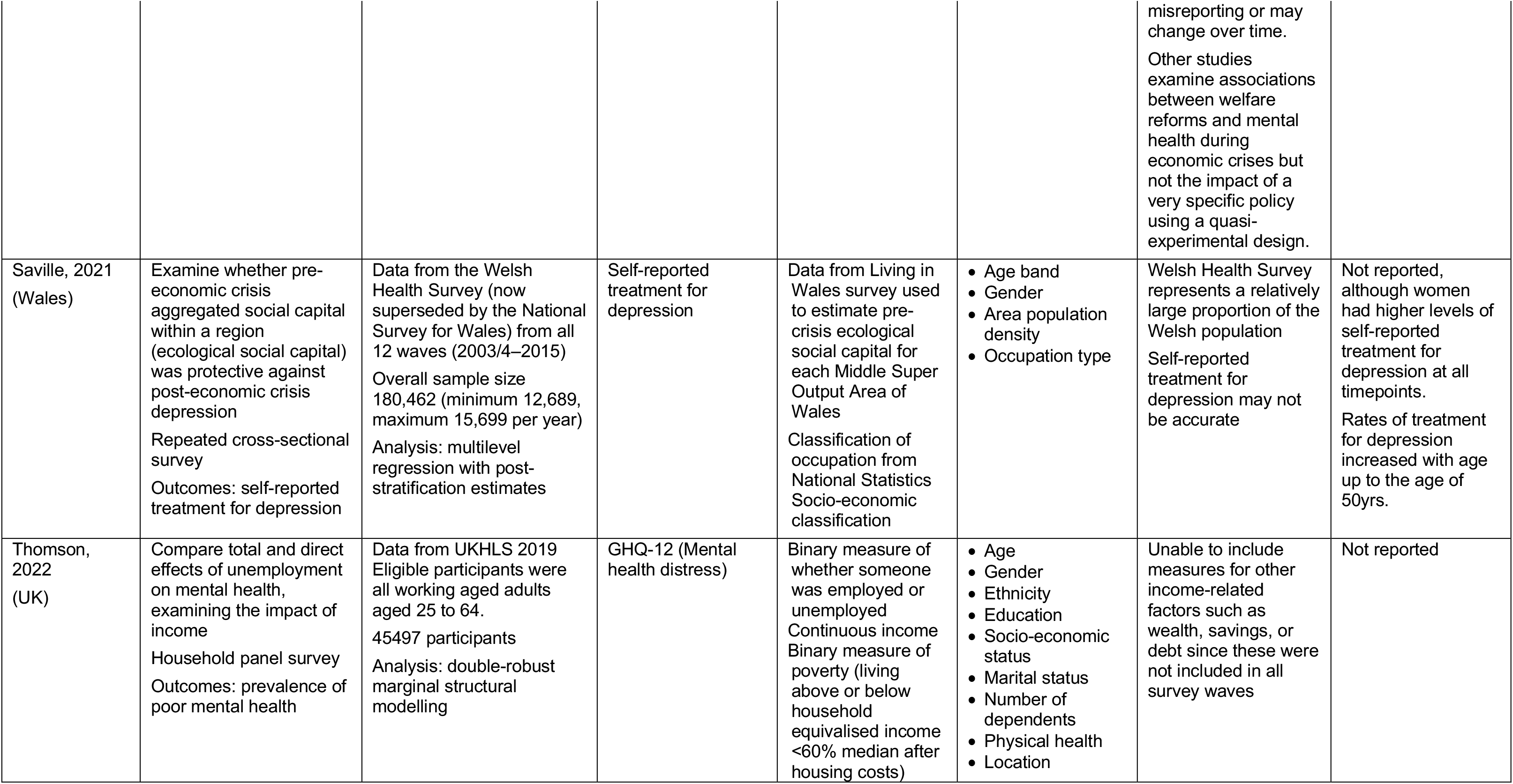
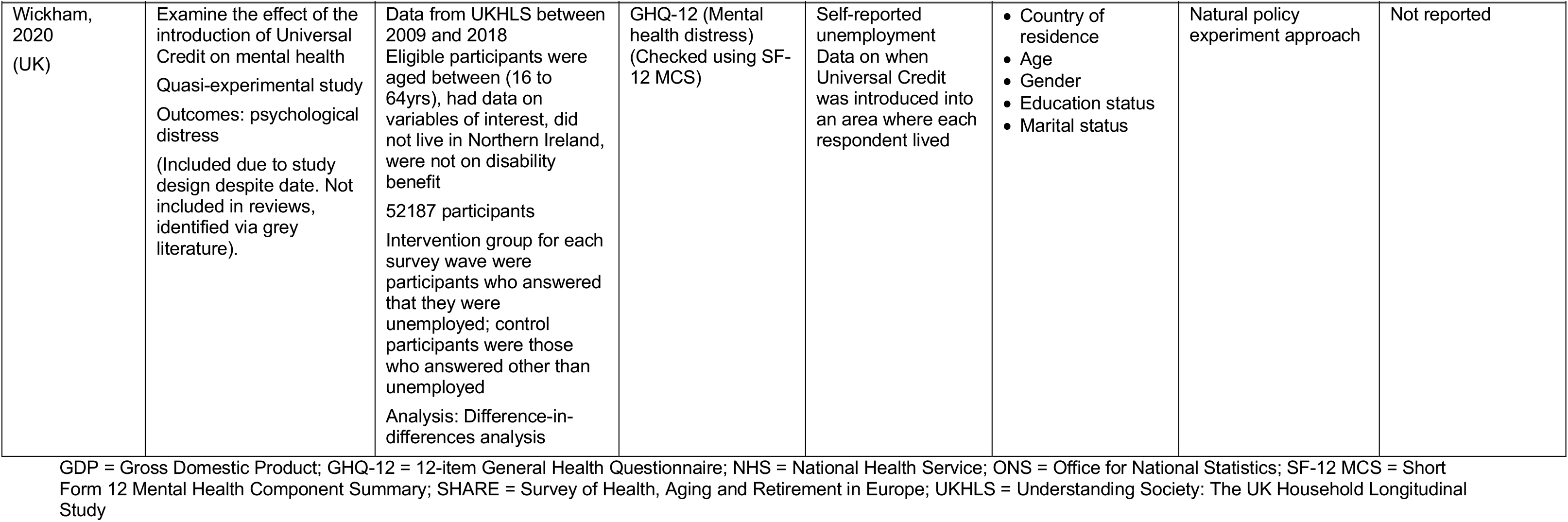
Primary studies.

| Citation (Country) | Study Details (purpose, design, mental health outcomes) | Study characteristics (source of data, details of sample, dates of data collection, analysis) | Mental health measurement tools and methods | Specific economic measures | Covariates / confounders | Author assessment of strengths and limitations of measurement | Groups identified as particularly vulnerable |
| --- | --- | --- | --- | --- | --- | --- | --- |
| Alvarez-Galvez, 2021 (Spain) | Assess association between recent economic downturns and age-standardised suicide rates adjusting for social cohesion and community values (interpersonal trust, social capital, post materialist values obtained from World Values Survey)<br>Time-series study<br>Outcomes: Suicide | Population-level data<br>Age-standardised suicide rates from:<br>(1) 1980 to 1991; (2) from 1992 until 2007; (3) from 2008 to 2010; (4) from 2011 to 2017<br>Interrupted time-series analysis | Suicide rates per 100,000 obtained from the National Statistics Institute of Spain | From Eurostat data:<br>• Unemployment rate<br>• GDP growth<br>• Social expenditure as a percentage of GDP | Social relationships data from the World Values Survey:<br>• Interpersonal trust<br>• Social capital<br>• Postmaterialist values<br>Men and women analysed separately<br>Age standardised results presented | Methodological variations in suicide data from registries.<br>Aggregate national data used which means individual behaviour cannot be inferred. | Men more at risk of suicide at all timepoints |
| Artez, 2022 (Netherlands/Germany) | Investigate whether the Great Recession (2007 to 2009) increased the risk of depression in older people in Europe.<br>Panel study<br>Outcomes: late-life depression | Data from Survey of Health, Ageing and Retirement in Europe (waves 1 – 7), conducted in Austria, Belgium, Denmark, France, Germany, Italy, Spain, Sweden, Switzerland<br>Data from 2004 to 2017<br>Eligible participants were ≥50yrs in wave 1; had valid depression data in waves 1 and 2 (prior to Great Repression) and for at least one wave after; depression-free at baseline; did not relocate during study period | EURO-D depression symptom scale | Country-level GDP data used to identify start of recession by country<br>Perceived area deprivation (from SHARE) | Individual level confounders:<br>• Age<br>• Job status<br>• Physical activity<br>• Cognitive functioning<br>• Frailty<br>• Activity of daily living<br>• Number of chronic diseases<br>Household level confounders:<br>• Living together with a partner<br>• Household size<br>• Urban/rural | EURO-D was developed to specifically to measure late-life depression<br>Use of a mental health questionnaire identifies undiagnosed depression<br>Survey only conducted every 2 years, which limits ability to identify very short-term effects<br>Individual-level confounders available<br>(Note: This study is useful to show how a | People living in more deprived areas |
|  |  | 6866 participants eligible at baseline (out of 95534); 3144 included in fixed effects model (53% female)<br>Analysis: Binary logistic individual fixed effect (FE) models using within-estimators. |  |  |  | subset from cohort data can be used/derived). |  |
| Cherrie, 2021 (Scotland) | Examine how the trend in 'new' antidepressant prescription rates during the Great Recession varied by regional economic conditions<br>Longitudinal data linkage study<br>Outcomes: Anti-depressant prescriptions | Data from the Scottish Longitudinal Study, linked to NHS service use and prescriptions for mental health conditions<br>Eligible participants supplied data in 2001 and 2011 censuses (pre- and post- recession onset); were aged between 16 and 60 in 2011; were economically active and employed in the labour market in 2011.<br>86500 participants eligible<br>Linked to NHS data from 2009 to 2014<br>Analysis: Multilevel logistic regression and mediation analysis | New anti-depressant prescriptions from linked NHS data | Labour market trajectories derived from annual trends in full-time employment (derived from NOMIS Official Labour Market Statistics, ONS) from 2004 to 2014<br>Average regional income lost per working adult due to welfare reforms | • Age<br>• Gender<br>• Ethnicity<br>• Marital status<br>• Living alone<br>• Social grade<br>• Carstairs neighborhood deprivation<br>• Health selective migration prior to study period controlled for with data on Local Authority of residence in 2001 and 2011 | Population representative sample used<br>Linked data on prescribing<br>No data on diagnosis of depression so some prescriptions may not be for mental illness<br>Unable to control for differences between prescribers in treating depression with anti-depressants | People living in areas with low or declining levels of full-time employment |
| Clair, 2022 (UK) | Explore the impact of cold homes on mental health, the role of gender as a mediator and the effect of transitioning into a cold home<br>Household panel study | Data from UKHLS 2009 to Dec 2021<br>Eligible participants entered the study at wave 1; ≥16yrs; had data for at least 3 waves<br>Sample stratified into: | GHQ-12 (Mental health distress) (Checked with SF-12 MCS) | Cold home measured by responses to question: "In winter are you able to keep this accommodation warm enough" | • Age<br>• Gender<br>• Ethnicity<br>• Region<br>• Employment status | Observed that it is possible that mental distress perceptions of temperature; that different people may experience household | People transitioning into living in cold homes |
|  | Outcomes: mental distress<br>Note: During the study period energy prices, housing costs were rising | Good mental health and lived in warm home at wave 1 (n=21281)<br>Borderline mental health and lived in warm home at wave 1 (n=2258)<br>Analysis: multilevel discrete-time event models |  |  | • Diagnosis of longstanding condition or disability<br>• Adjusted income quartile<br>• Highest educational qualification<br>• Household composition<br>• Financial situation<br>• Household tenure<br>• Building type<br>• Housing payment arrears | temperature differently |  |
| Curtis, 2021 (England) | Determine if the risk of worsening mental health was greater for people living in the most deprived areas in terms of unemployment and impact of austerity policies<br>Household panel survey<br>Outcomes: Trends in self-reported mental health | Data from UKHLS waves 3 to 7, 2011 to 2017<br>Eligible participants were ≥16yrs; living in England; had full data on the variables of interest.<br>17212 participants<br>Maximum likelihood multinomial logit models | SF-12 MCS (version 2) | Geographical variables to categorise place of residence according to socio-geographic aspects (employment domain of the Index of Multiple Deprivation; average income loss per person of working age due to welfare reforms; rural / urban; West Midlands/other)<br>Indicator showing whether someone had moved to a location in a different category | • Gender<br>• Age group<br>• Ethnicity<br>• Living with a partner<br>• Occupational social class<br>• Income in the month prior to interview<br>• Employment status<br>• Housing tenure at wave 4<br>• In receipt of welfare benefits at wave 4 (other than child benefits and state pensions) | Using a sub-sample from the panel survey means it may not no longer be representative of England as a whole | People who were more deprived<br>Living in economically disadvantaged neighborhoods<br>Living in areas most impacted by austerity policies<br>Becoming unemployed<br>Moving onto welfare benefits |
|  |  |  |  |  | • Change in socio-economic status |  |  |
| Kim, 2022 (England) | Examine the impact of the announcement (April 2012) and implementation (April 2013) of the spare room subsidy ("bedroom tax") on psychological distress in social housing renters<br>Quasi-experimental study with matched controls<br>Outcomes: psychological distress | Data from UKHLS 2010 to 2014 for main analysis and up to 2017 for sensitivity tests.<br>Eligible participants were living in social housing; had pre- and post- policy data; were over 16 and under state pension age; lived in England<br>2078 eligible participants, of which 422 were living in underoccupied housing and were affected by the policy prior to April 2013 ('treatment group') and 1926 were not affected.<br>Final matched sample = 412 treatment group and 412 controls.<br>Difference-in-differences analysis | GHQ-12 (Mental health distress) | None | • Observed and unobserved individual- level time-invariant characteristics (e.g., immigration status and family history of illnesses)<br>• Regional differences | Pre- and post- design and use of matching and appropriate analysis enhances ability to determine causality.<br>Other covariates not captured may confound the treatment effects of an intervention<br>GHQ-12 is limited compared with formal mental health assessment from a trained health care professional<br>Loss to follow-up is common in panel surveys, which may bias estimates if different between control and treatment arms.<br>Matching may reduce representativeness of the sample<br>Treatment status was assumed based on household characteristics prior to the intervention period; household characteristics may be vulnerable to<br>misreporting or may change over time.<br>Other studies examine associations between welfare reforms and mental health during economic crises but not the impact of a very specific policy using a quasi-experimental design. | People of lower socioeconomic status |
| Saville, 2021 (Wales) | Examine whether pre-economic crisis aggregated social capital within a region (ecological social capital) was protective against post-economic crisis depression<br>Repeated cross-sectional survey<br>Outcomes: self-reported treatment for depression | Data from the Welsh Health Survey (now superseded by the National Survey for Wales) from all 12 waves (2003/4–2015)<br>Overall sample size 180,462 (minimum 12,689, maximum 15,699 per year)<br>Analysis: multilevel regression with post-stratification estimates | Self-reported treatment for depression | Data from Living in Wales survey used to estimate pre-crisis ecological social capital for each Middle Super Output Area of Wales<br>Classification of occupation from National Statistics Socio-economic classification | • Age band<br>• Gender<br>• Area population density<br>• Occupation type | Welsh Health Survey represents a relatively large proportion of the Welsh population<br>Self-reported treatment for depression may not be accurate | Not reported, although women had higher levels of self-reported treatment for depression at all timepoints.<br>Rates of treatment for depression increased with age up to the age of 50yrs. |
| Thomson, 2022 (UK) | Compare total and direct effects of unemployment on mental health, examining the impact of income<br>Household panel survey<br>Outcomes: prevalence of poor mental health | Data from UKHLS 2019<br>Eligible participants were all working aged adults aged 25 to 64.<br>45497 participants<br>Analysis: double-robust marginal structural modelling | GHQ-12 (Mental health distress) | Binary measure of whether someone was employed or unemployed<br>Continuous income<br>Binary measure of poverty (living above or below household equivalised income <60% median after housing costs) | • Age<br>• Gender<br>• Ethnicity<br>• Education<br>• Socio-economic status<br>• Marital status<br>• Number of dependents<br>• Physical health<br>• Location | Unable to include measures for other income-related factors such as wealth, savings, or debt since these were not included in all survey waves | Not reported |
| Wickham, 2020 (UK) | Examine the effect of the introduction of Universal Credit on mental health<br>Quasi-experimental study<br>Outcomes: psychological distress<br>(Included due to study design despite date. Not included in reviews, identified via grey literature). | Data from UKHLS between 2009 and 2018<br>Eligible participants were aged between (16 to 64yrs), had data on variables of interest, did not live in Northern Ireland, were not on disability benefit<br>52187 participants<br>Intervention group for each survey wave were participants who answered that they were unemployed; control participants were those who answered other than unemployed<br>Analysis: Difference-in-differences analysis | GHQ-12 (Mental health distress)<br>(Checked using SF-12 MCS) | Self-reported unemployment<br>Data on when Universal Credit was introduced into an area where each respondent lived | Country of residence<br>Age<br>Gender<br>Education status<br>Marital status | Natural policy experiment approach | Not reported |

GDP = Gross Domestic Product; GHQ-12 = 12-item General Health Questionnaire; NHS = National Health Service; ONS = Office for National Statistics; SF-12 MCS = Short Form 12 Mental Health Component Summary; SHARE = Survey of Health, Aging and Retirement in Europe; UKHLS = Understanding Society: The UK Household Longitudinal Study

**Table 3c:** Summary of grey literature.

| Citation (country) | Details (aim, study type, organisation) | Mental health measurement tools and methods | Specific economic measures | Author assessment of methods | Groups identified as particularly vulnerable |
| --- | --- | --- | --- | --- | --- |
| Clark, 2022 (UK) | Report examining associations between insecurity and anxiety in the UK<br>Uses data from GP records, national surveys (panel surveys and repeated cross-sectional), although some data used is cross-sectional being taken from a single year.<br>Joseph Rowntree Foundation | • GHQ -12<br>• SF-12<br>• Warwick-Edinburgh mental well-being scale (WEMWBS)<sup>1</sup><br>• Single anxiety score<br>Medical records:<br>• GP recording diagnosis of anxiety from: Clinical Practice Research Datalink<br>• The Health Improvement Network<br>Population survey:<br>• UKHLS<br>• Annual Population Survey | Employment data:<br>• Office for National Statistics<br>• UKHLS<br>• Annual Population Survey<br>Housing data:<br>• Labour Force Survey<br>• English Housing Survey<br>Household savings:<br>• Wealth and Assets Survey<br>• Family Resources Survey<br>Social capital:<br>• Office for National Statistics<br>Deprivation:<br>• Index of Multiple Deprivation | Prescribing data needs care with interpretation since the availability of drugs may vary and different prescribers may have different prescribing thresholds and the same drug may be given for different conditions<br>Noted that UK official surveys do not always ask about both mental health and household economics in detail, may miss specific economic details and data from different surveys cannot be linked.<br>The authors report that UKHLS has 10,000s of respondents and has information on both health and economics, although there are difficulties in identifying people in insecure work or forced freelancers.<br>UKHLS allows individuals to be tracked over time to determine how different events affect mental health.<br>Overall, the authors report that official statistics and secondary sources on mental health and financial stress are easily available in the UK. | Living in debt<br>Living in rental accommodation (especially social renters)<br>Less than £1000 in savings or expecting less than enough as a pension |
| Preece 2019 (Wales) | Policy report / rapid review<br>Examine the relationship between housing insecurity and mental health, identify key lessons and relate to Wales<br>Post 2008 economic crisis | Mental health measurement tools:<br>• GHQ-12 (Mental health distress)<br>Population survey:<br>• UKHLS | Landlord possession rates and orders<br>Mortgage repossession rates | Although UKHLS data has been used there is limited data on housing characteristics, and it was not possible to control for housing condition. | Households with dependent children<br>People receiving means tested benefits<br>Migrant groups |
|  | Rapid review: literature searched from 2008 – 2019.<br>39 studies identified from searches, plus an unspecified number from handsearching<br>UK Collaborative Centre for Housing Evidence |  |  | Authors note that data gaps around housing and mental health is not drawn specifically from Wales (evidence is mainly from Australia and England) and may not be generalisable. | People experiencing socio-economic deprivation<br>People living with existing mental health problems |
| Office for National Statistics, 2022 (Great Britain) | To examine depression in the context of rising cost-of-living<br>Brief report describing repeated cross-sectional data collected fortnightly<br>Office for National Statistics | Mental health measurement tool: PHQ-8 (Depression)<br>Population survey: The Opinions and Lifestyle Survey | Specific questions asked during survey on e.g., housing tenure, ease of paying bills, savings etc. | Authors make a point of stating that no causal link between rising cost-of-living and depression is suggested | • Economically inactive because of long-term sickness<br>• Unpaid carers for 35 or more hours a week<br>• Disabled adults<br>• Living in the most deprived areas of England<br>• Young adults aged 16 to 29 years<br>• Single person household<br>• Women<br>• Living in rental accommodation |

GHQ-12 = 12-item General Health Questionnaire; PQH-8 = 8-item Public Health Questionnaire; SF-12 = Short Form 12; UKHLS = Understanding Society: The UK Household Longitudinal Study
<sup>1</sup>Also used in UKHLS, although data from this questionnaire was not utilised in this study.

#### 6.2.2 Mental Health Measurement Scales

**Table 4.**
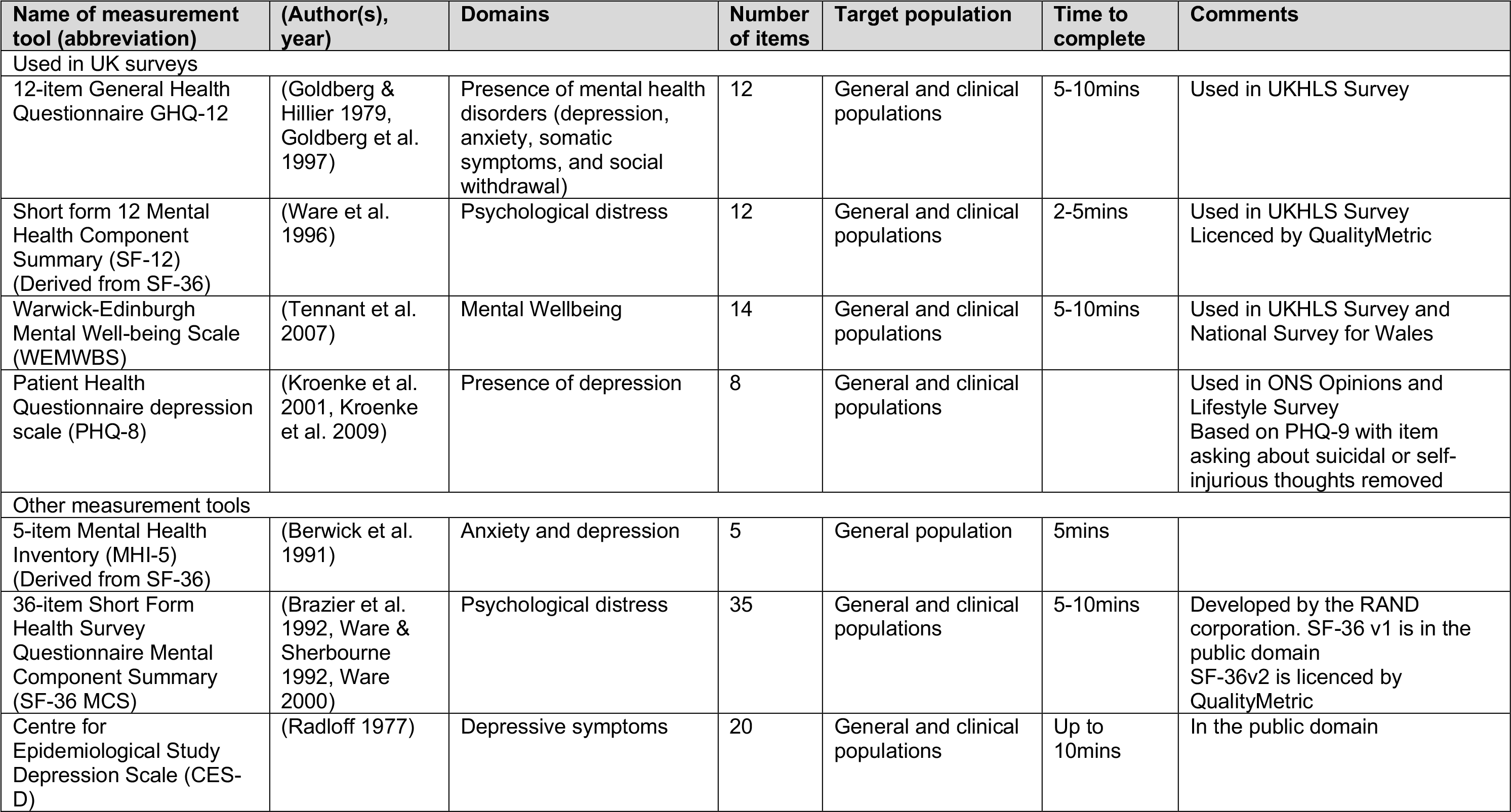
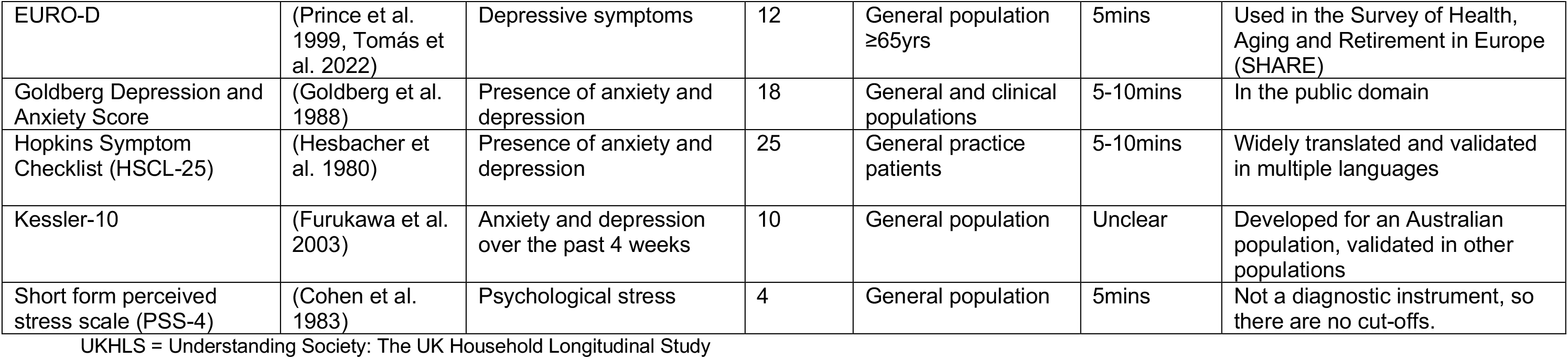
Mental Health Measurement Scales.

| Name of measurement tool (abbreviation) | (Author(s), year) | Domains | Number of items | Target population | Time to complete | Comments |
| --- | --- | --- | --- | --- | --- | --- |
| Used in UK surveys |  |  |  |  |  |  |
| 12-item General Health Questionnaire GHQ-12 | (Goldberg & Hillier 1979, Goldberg et al. 1997) | Presence of mental health disorders (depression, anxiety, somatic symptoms, and social withdrawal) | 12 | General and clinical populations | 5-10mins | Used in UKHLS Survey |
| Short form 12 Mental Health Component Summary (SF-12) (Derived from SF-36) | (Ware et al. 1996) | Psychological distress | 12 | General and clinical populations | 2-5mins | Used in UKHLS Survey<br>Licenced by QualityMetric |
| Warwick-Edinburgh Mental Well-being Scale (WEMWBS) | (Tennant et al. 2007) | Mental Wellbeing | 14 | General and clinical populations | 5-10mins | Used in UKHLS Survey and National Survey for Wales |
| Patient Health Questionnaire depression scale (PHQ-8) | (Kroenke et al. 2001, Kroenke et al. 2009) | Presence of depression | 8 | General and clinical populations |  | Used in ONS Opinions and Lifestyle Survey<br>Based on PHQ-9 with item asking about suicidal or self-injurious thoughts removed |
| Other measurement tools |  |  |  |  |  |  |
| 5-item Mental Health Inventory (MHI-5) (Derived from SF-36) | (Berwick et al. 1991) | Anxiety and depression | 5 | General population | 5mins |  |
| 36-item Short Form Health Survey Questionnaire Mental Component Summary (SF-36 MCS) | (Brazier et al. 1992, Ware & Sherbourne 1992, Ware 2000) | Psychological distress | 35 | General and clinical populations | 5-10mins | Developed by the RAND corporation. SF-36 v1 is in the public domain<br>SF-36v2 is licenced by QualityMetric |
| Centre for Epidemiological Study Depression Scale (CES-D) | (Radloff 1977) | Depressive symptoms | 20 | General and clinical populations | Up to 10mins | In the public domain |
| EURO-D | (Prince et al. 1999, Tomás et al. 2022) | Depressive symptoms | 12 | General population ≥65yrs | 5mins | Used in the Survey of Health, Aging and Retirement in Europe (SHARE) |
| Goldberg Depression and Anxiety Score | (Goldberg et al. 1988) | Presence of anxiety and depression | 18 | General and clinical populations | 5-10mins | In the public domain |
| Hopkins Symptom Checklist (HSCL-25) | (Hesbacher et al. 1980) | Presence of anxiety and depression | 25 | General practice patients | 5-10mins | Widely translated and validated in multiple languages |
| Kessler-10 | (Furukawa et al. 2003) | Anxiety and depression over the past 4 weeks | 10 | General population | Unclear | Developed for an Australian population, validated in other populations |
| Short form perceived stress scale (PSS-4) | (Cohen et al. 1983) | Psychological stress | 4 | General population | 5mins | Not a diagnostic instrument, so there are no cut-offs. |

UKHLS = Understanding Society: The UK Household Longitudinal Study

### 6.3 Information available on request

The study protocol, full search strategy and list of excluded studies are available upon request.

## 7 ADDITIONAL INFORMATION

### 7.1 Conflicts of interest

The authors declare they have no conflicts of interest to report.

## 7.2 Acknowledgements

The authors would like to thank James Burgess, Brendan Collins, Rebecca Masters, and Nigel Pearson for their time and contributions during stakeholder meetings in guiding the focus of the review and interpretation of findings.

## APPENDIX 1: Resources searched during Rapid Review Searching

**Table A1.1.**
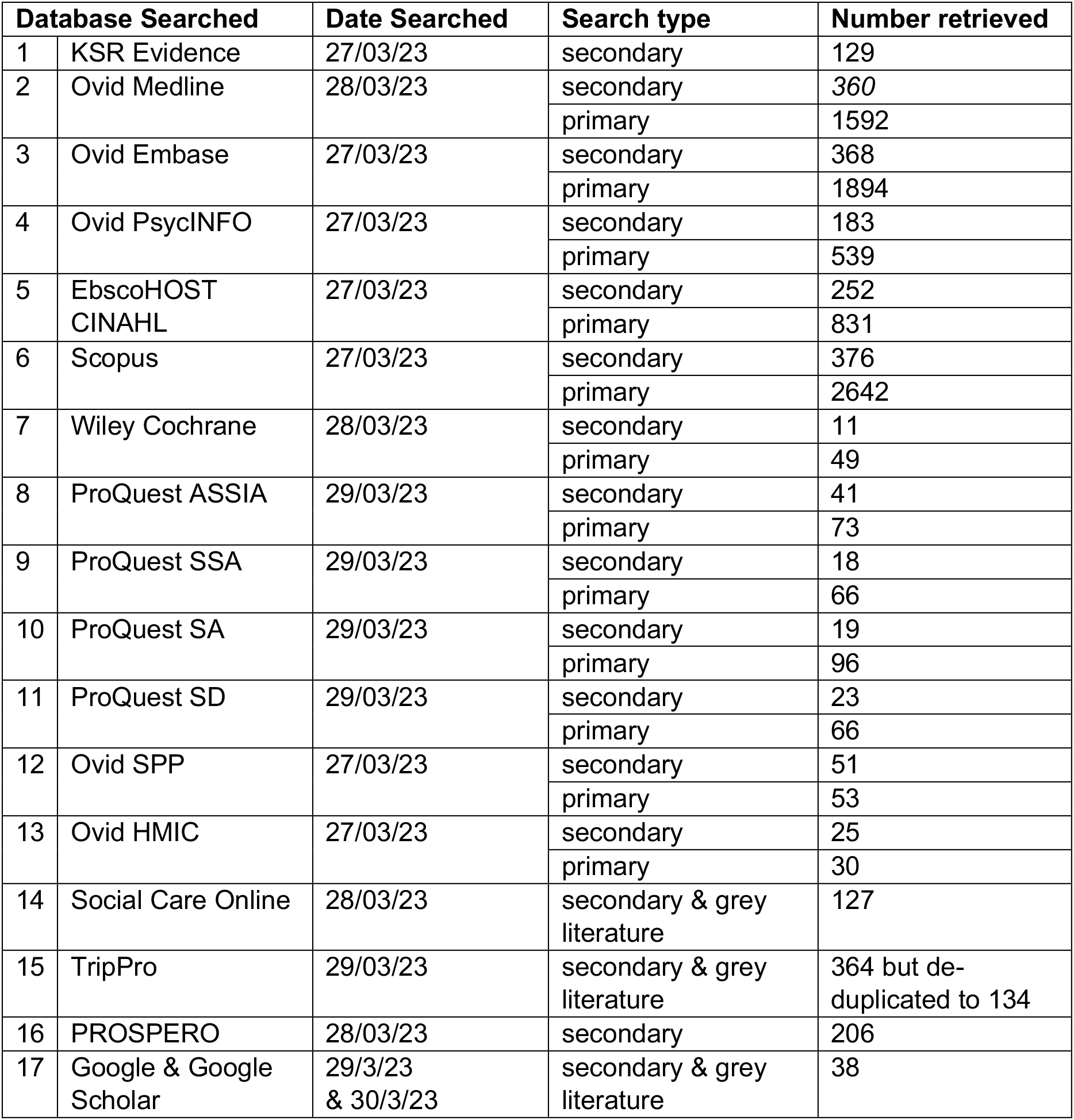
Main evidence searches.

**Table A1.2.**
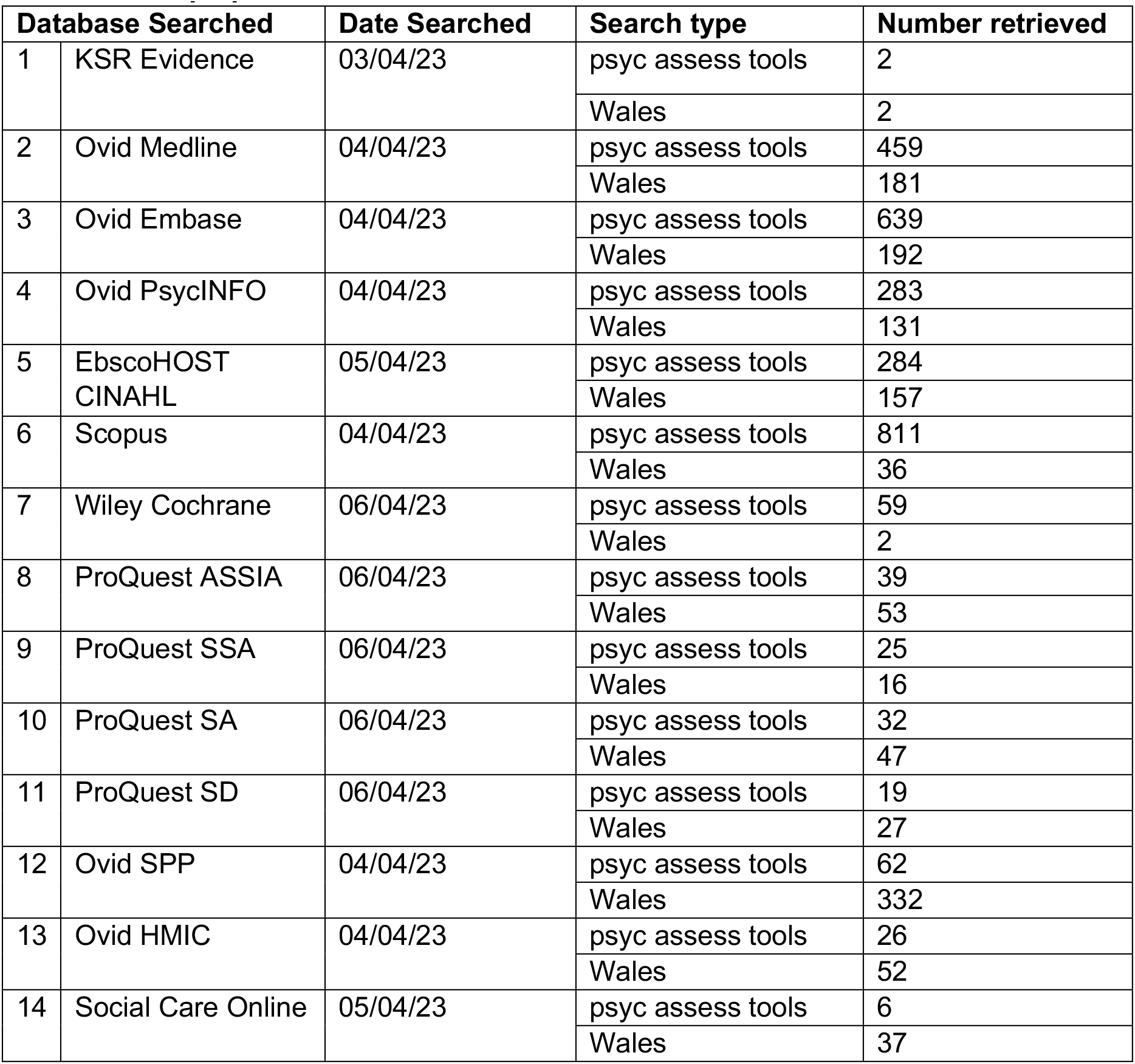
Mop up searches.

## APPENDIX 2: MEDLINE STRATEGY

**Table A2.1:**
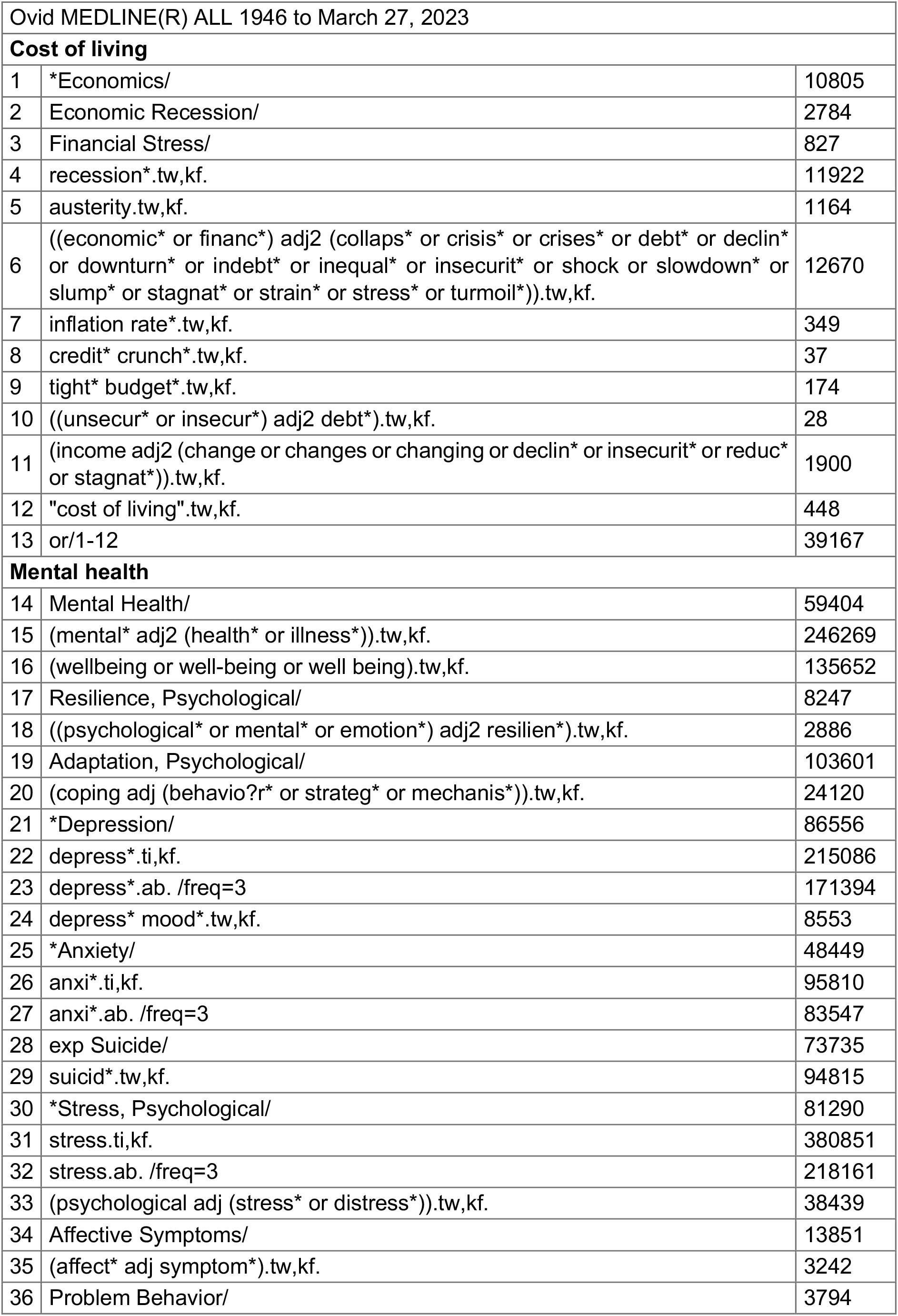
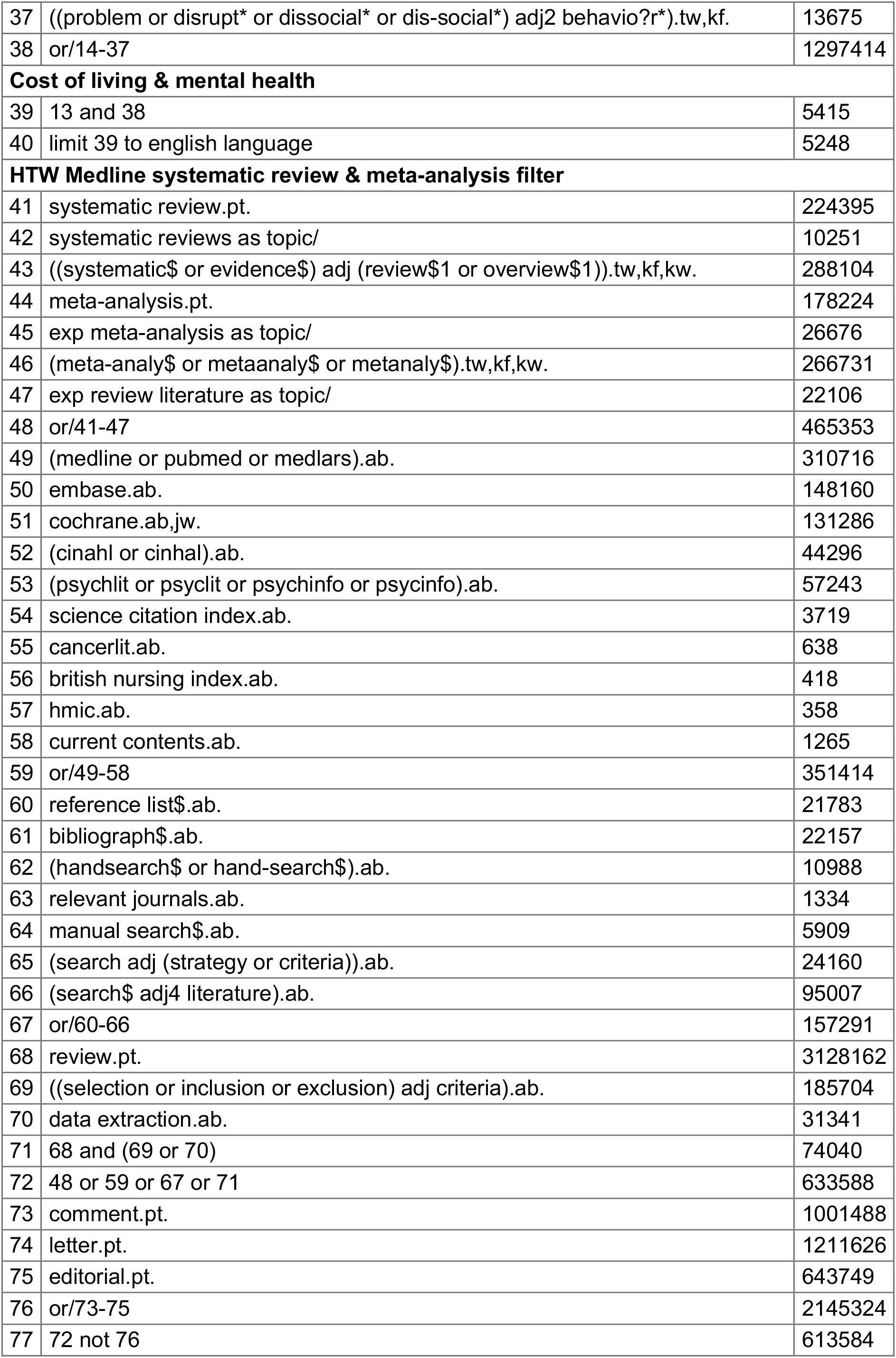
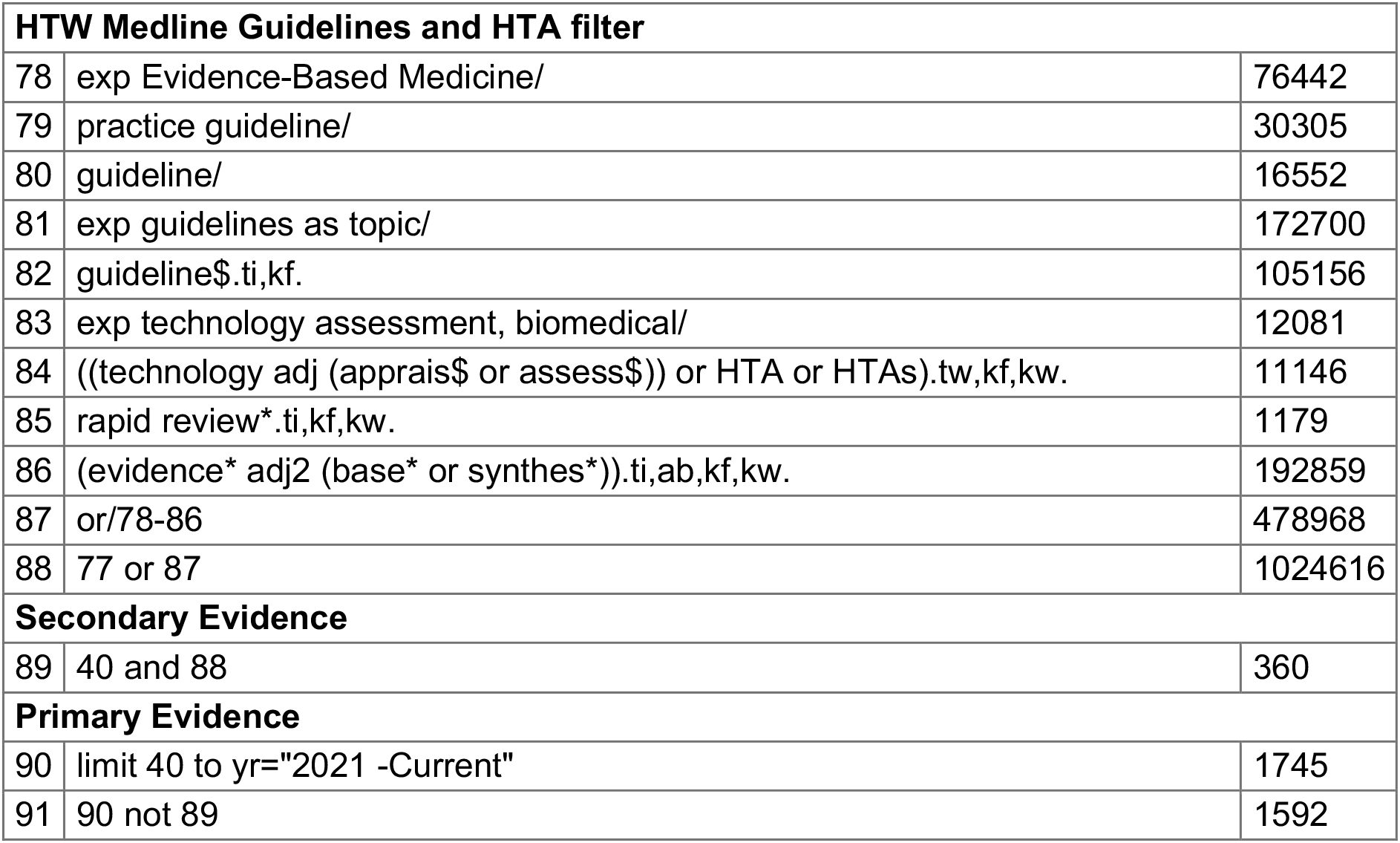
Main searches.

**Table A2.2.**
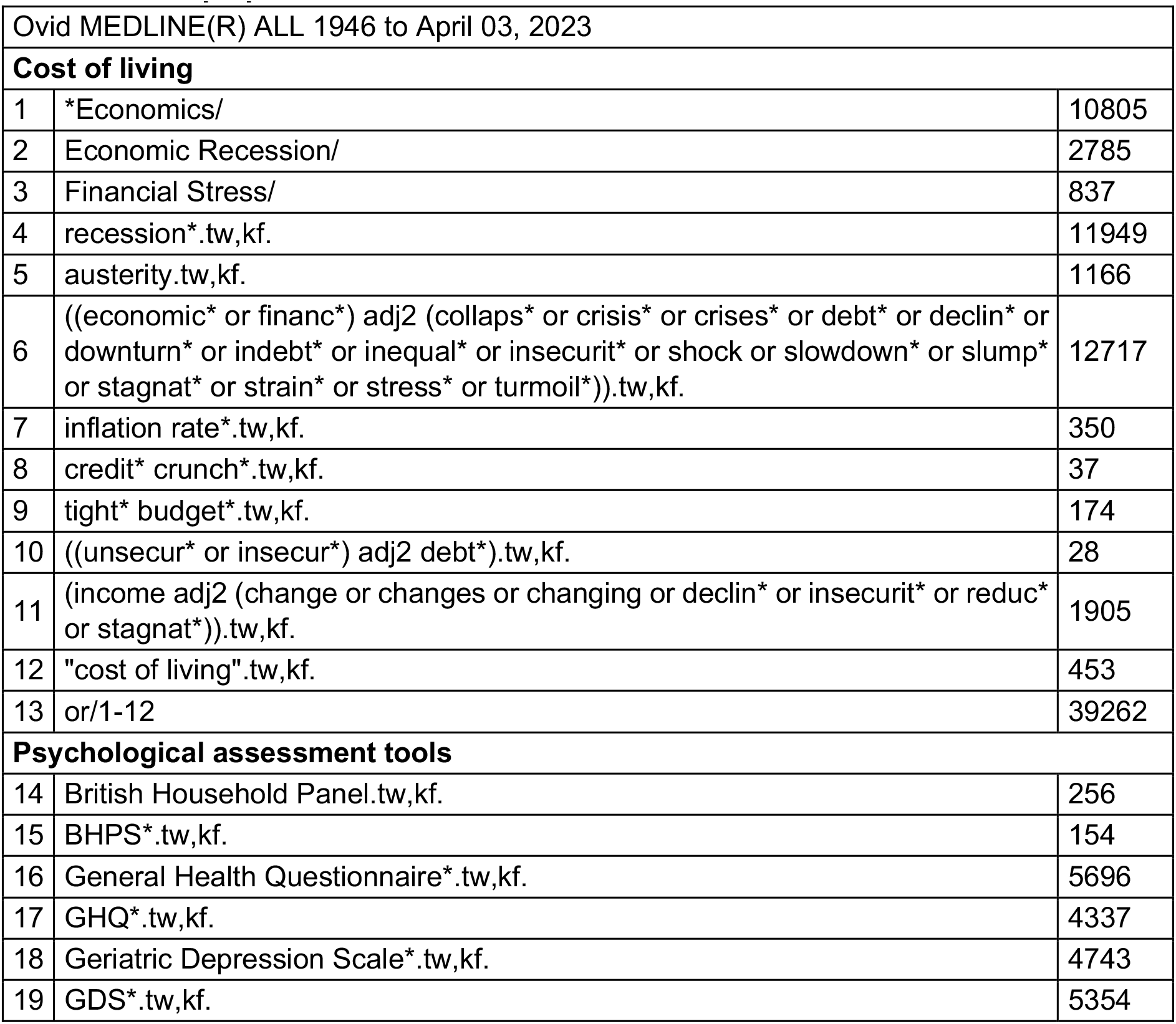
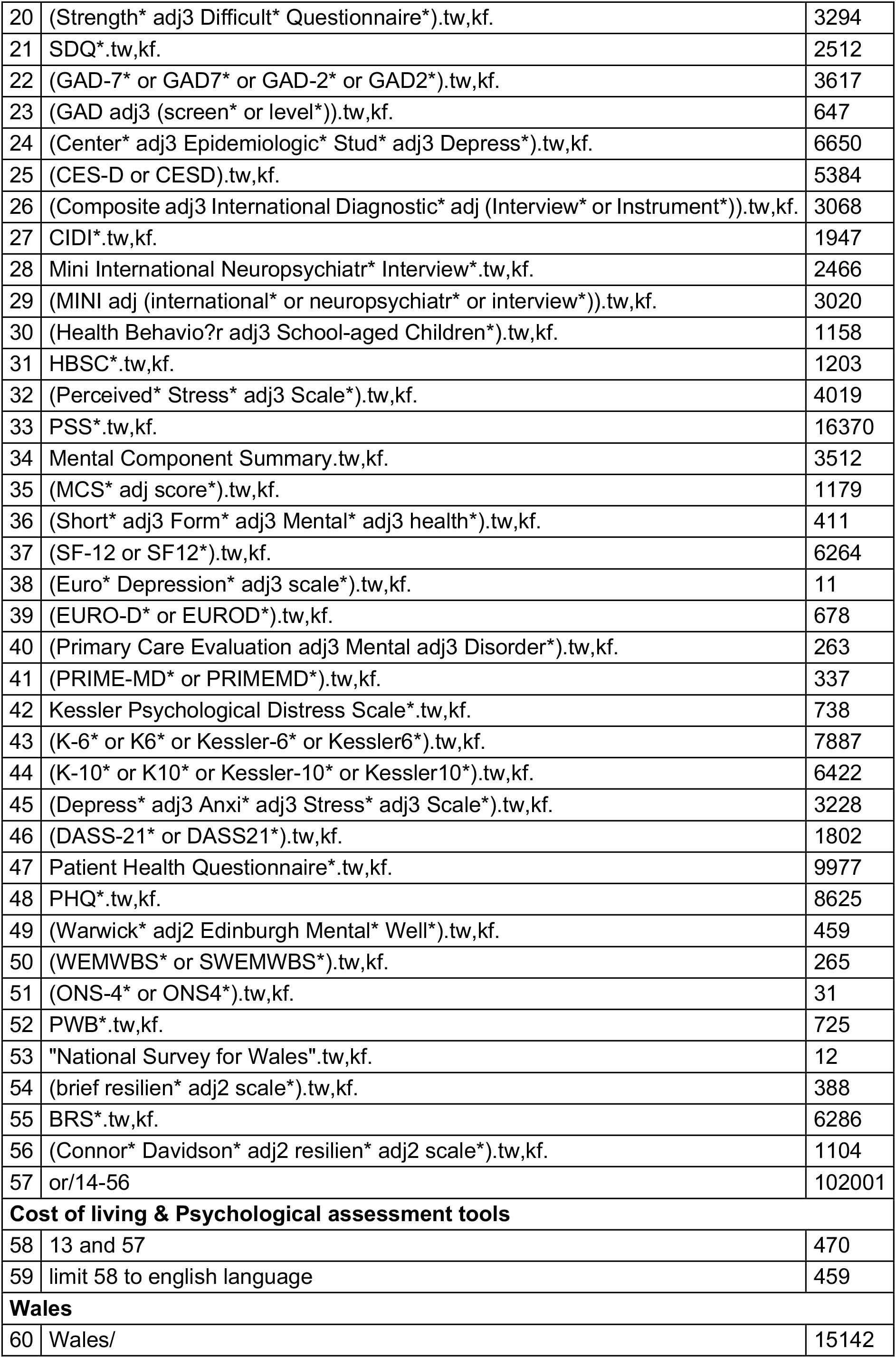
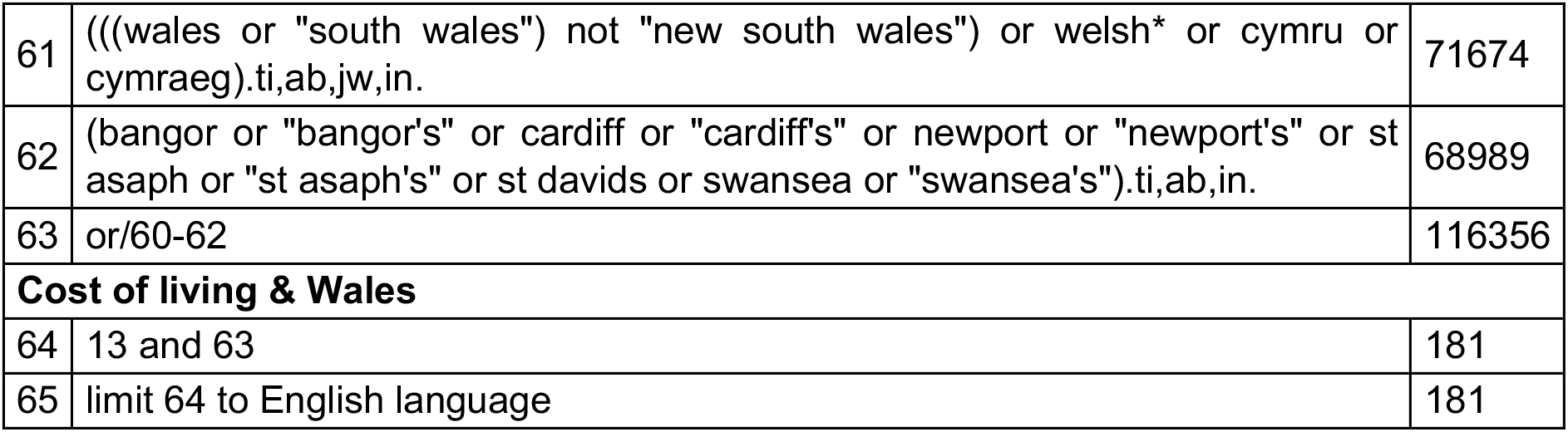
Mop up searches.

## Footnotes

* This section has been completed by the Centre for Health Economics & Medicines Evaluation (CHEME), Bangor University

## Notes

### Competing Interest Statement

The authors have declared no competing interest.

